# Benefits of natural habitat particularly woodland on children’s cognition and mental health

**DOI:** 10.1101/2021.01.12.21249675

**Authors:** Mikaël J. A. Maes, Monica Pirani, Elizabeth R. Booth, Chen Shen, Ben Milligan, Kate E. Jones, Mireille B. Toledano

## Abstract

Life in urban areas is associated with adverse human health effects, including risks of developing cognitive problems and mental health issues. Many epidemiological studies have established associations between urban nature, cognitive development and mental health, but why specifically we receive these health benefits remains unclear, especially in children. Here, we used longitudinal data in a cohort of 3,568 children aged 9 to 15 years at 31 schools across London to develop a model and examine the associations between natural habitat type, and children’s cognitive development and mental health. We show that, after adjusting for other environmental, demographic and socioeconomic variables, higher daily exposure rates to natural habitat and particularly woodland were associated with enhanced cognitive development and mental health from late childhood to early adolescence. Our results suggest that optimising ecosystem services linked to cognitive development and mental health benefits should prioritise the type of natural habitat for sustainable urban planning decisions.

## INTRODUCTION

Over 55% of the human population is now living in cities, with an estimated 2.5 billion more people to be added to the urban population by 2050^1^. Although urban populations often have better socioeconomic livelihoods, living in urban areas is associated with a number of adverse human health effects^2^. In particular, urbanisation is associated with risks of developing cognitive problems and mental health issues^3,4^, and is linked with various demographic and socioeconomic factors^5,6^. The COVID-19 pandemic has further exacerbated mental health problems such as stress, anxiety and depressive symptoms, amongst others^7,8^. In London, for example, an estimated 1 in 4 individuals will experience a diagnosable mental health condition in any given year, costing £26 billion annually through poorer education, employment and quality of life, affecting London’s economy, population and infrastructure^9^. The renewed focus on cognition and mental health due to the negative effects of the COVID-19 pandemic highlight the importance to understand the mechanisms and dynamic interactions attributed to a higher risk of cognitive problems and mental health issues in urban areas, which until now remain unclear.

Emerging evidence suggests that surrounding environments, particularly exposure to natural areas plays an important role in cognitive development and mental health^10–12^, and are part of a range of ecosystem services (ES) characterised to impact human health and well-being^13^. Relative risk estimates of the association between natural areas and cognitive development and mental health have been comparable in magnitude to family history and parental age, and higher than the degree of urbanisation^11^. However, these associations lack a mechanistic understanding^14^.

Sensory and non-sensory pathways have been suggested as potentially important to deliver health benefits received from nature^14^. Although humans are multisensory, previous studies have primarily focused on the visual sense in humans^15,16^. However, natural sounds and smells, for example, are preferred over anthropogenic ones^17,18^, suggesting that non-visual psychological pathways are also important. Health benefits received from inhalation or ingestion of phytoncides, negative air ions or microbes may well explain other non-sensory physiological pathways through which health benefits are received from nature^19,20^. Further research into sensory and non-sensory pathways might prove fundamentally important to establish a mechanistic pathway between nature and mental health.

One of the barriers to understanding the mechanistic relationships between nature, cognitive development and mental health is the use of inconsistent exposure definitions in previous studies. Nature exposure has been measured, amongst others, as physical access to nature^21^, natural habitat type^22,23^, nature dose^24^ and degree of urbanisation^11,24^. Wider-scale epidemiological research studying the association between nature and mental health has almost exclusively measured ‘greenness’ through vegetation indices such as the Normalized Difference Vegetation Index (NDVI), a unit-less index of relative overall vegetation density and quality^10–12,25^. However, this tends to simplify ‘greenness’ without taking into account the types of natural habitat that exist. For example, standing and flowing water bodies such as lakes, rivers or reservoirs (hereinafter called blue space), are often excluded from nature and mental health research^10,11,26^, and when included, show significant associations with mental health and cognitive development^25,27^. In addition, forest has been proposed to generate a more restorative effect both psychologically^23,26^ and physiologically^19^. For example, studies have shown that forests have a more restorative effect when compared with overall urban green space, agricultural land or wetland, amongst others^23,26^. Although many of these measures to assess environmental exposure have found associations with mental health or cognitive development, to date there is no comprehensive analysis or agreement which measure of environmental exposure is more or less important. A more nuanced understanding of these patterns would help to better understand the mechanisms behind such associations.

Many previous studies have also often focused on adult assessment of a variety of environmental exposures in relation to mental health and cognitive development^28^. However, there is growing recognition of the importance of children’s cognitive development and mental health, who are in the midst of their cognitive and mental development, affecting the child, its family and the broader community^29^. In fact, 1 in 10 of London’s children (∼111,600 children) between the ages of 5 and 16 suffer from a clinical mental health illness, and excess costs of mental health problems in children are estimated between £11,030 and £59,130 annually for each child^29^. As for adults, there is also evidence that surrounding nature plays an important role in children’s cognitive development and mental health into adulthood^11,12,25^. However, it remains unclear if the type of natural habitat influences the relationship with children’s cognitive development and mental health, nor what the relative impact of surrounding nature is with regards to other risk factors.

In this study, we develop a set of models estimating the contribution of urban natural habitat types to children’s cognitive development and mental health and argue these can be used to inform future urban planning decisions. Our models demonstrated that urban natural habitat and particularly woodland habitat mitigate cognitive problems and mental health issues in children from late childhood to early adolescence. We focused our analysis on a longitudinal dataset of 3,568 children from the Study of Cognition, Adolescents and Mobile Phones (SCAMP) across the London metropolitan area in the United Kingdom (Fig. 1a) (see Methods). We assessed cognitive development through a composite executive function (EF) score using computerised tests (Fig. 1b), while we assessed mental health through self-reported questionnaires on emotional and behavioural problems using the Strength and Difficulties Questionnaire (SDQ) total difficulties score (Fig. 1c), and overall well-being using the KIDSCREEN-10 Questionnaire Health-Related Quality of Life (HRQoL) score (Fig. 1d). We systematically mapped urban natural habitat to identify each child’s daily exposure rate (DER) around their home and school within 50 m, 100 m, 250 m and 500 m in a three-tier stepwise characterisation of urban natural habitat: (1) natural space, (2) green and blue space, and (3) grassland and woodland. Our models identified an important risk factor for children’s cognitive development and mental health and we suggest that this assists urban planners and decision-makers to sustainably manage urban nature. Unless stated otherwise, our results were based on fully adjusted models with natural habitat DERs with a daytime weighting and measured in buffer areas of 250 m (see Methods).

**Fig. 1.**
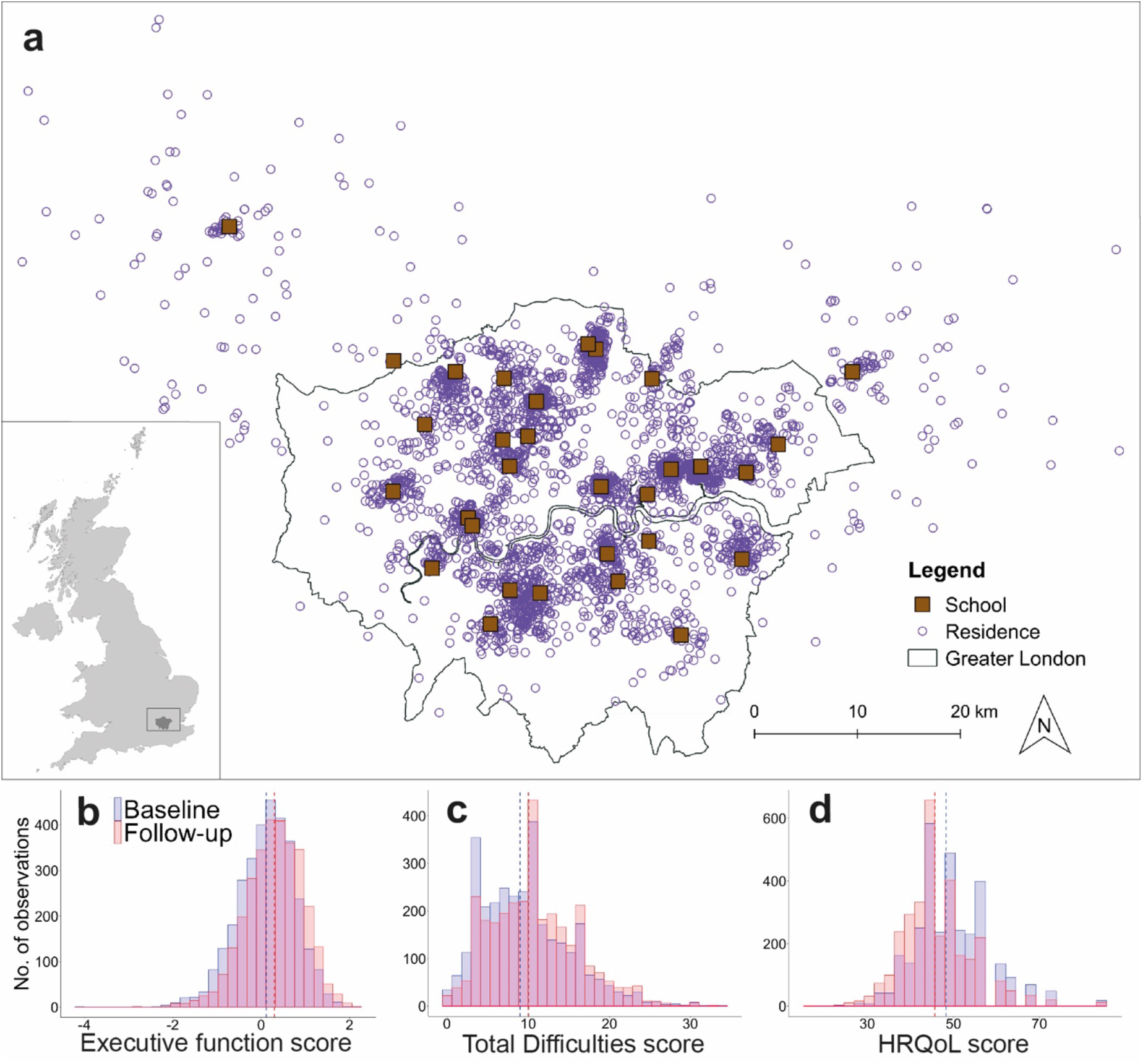
Geographic distribution of our longitudinal dataset and associated health variables for cognitive development and mental health. (a) Geolocation of the 3,568 children with a known home address during the baseline and follow-up assessment of the Study of Cognition, Adolescents and Mobile Phones and the 31 participating schools across the London metropolitan area, United Kingdom. Histograms show our baseline (blue) and follow-up (red) outcome for cognitive development: (b) Executive function score, and our outcomes for mental health: (c) Strengths and Difficulties Questionnaire total difficulties score and (d) KIDSCREEN-10 Questionnaire Health-Related Quality of Life score. A dashed line marks the median (Q_1_-Q_3_) for our baseline and follow-up outcomes, i.e. for (b) baseline: 0.16 (−0.30, 0.56), follow-up: 0.33 (−0.10, 0.76), (c) baseline: 9 (6, 13), follow-up: 10 (7, 14) and (d) baseline: 48.28 (43.34, 53.10), follow-up: 45.66 (41.23, 49.76).

## RESULTS

### Influence of urban natural habitat types for cognition and mental health

We estimated the change in children’s cognitive development and mental health for each type of urban natural habitat by fitting our longitudinal models (Supplementary Methods 1). We found that children’s cognitive development did improve with higher DER to natural space. A difference of one in natural space DER corresponded to an expected positive difference of 0.03 (95% credible interval [CI]: 0.01, 0.06) points in the child’s cognitive development using the EF score when controlling for other demographic, environmental and socio-economic fixed effects (Fig. 2a and Supplementary Figure 1a). We also provide the results for our mental health outcomes with natural space DER (Fig. 2b,c and Supplementary Figure 1b,c), where we found no improvement of mental health with higher DER to natural space, meaning the 95% CI included the null effect for both models. Our results for the tier 2 models, where both green and blue space was used for the natural habitat characterisation were almost identical to our tier 1 models for natural space DER. This is probably due to a high collinearity between our DER for natural space and green space (Supplementary Table 1) as children’s DER to blue space was low. This also meant that our models did not find an improvement of children’s cognitive development and mental health with DER of blue space (Fig. 2 and Supplementary Figure 2).

**Fig. 2.**
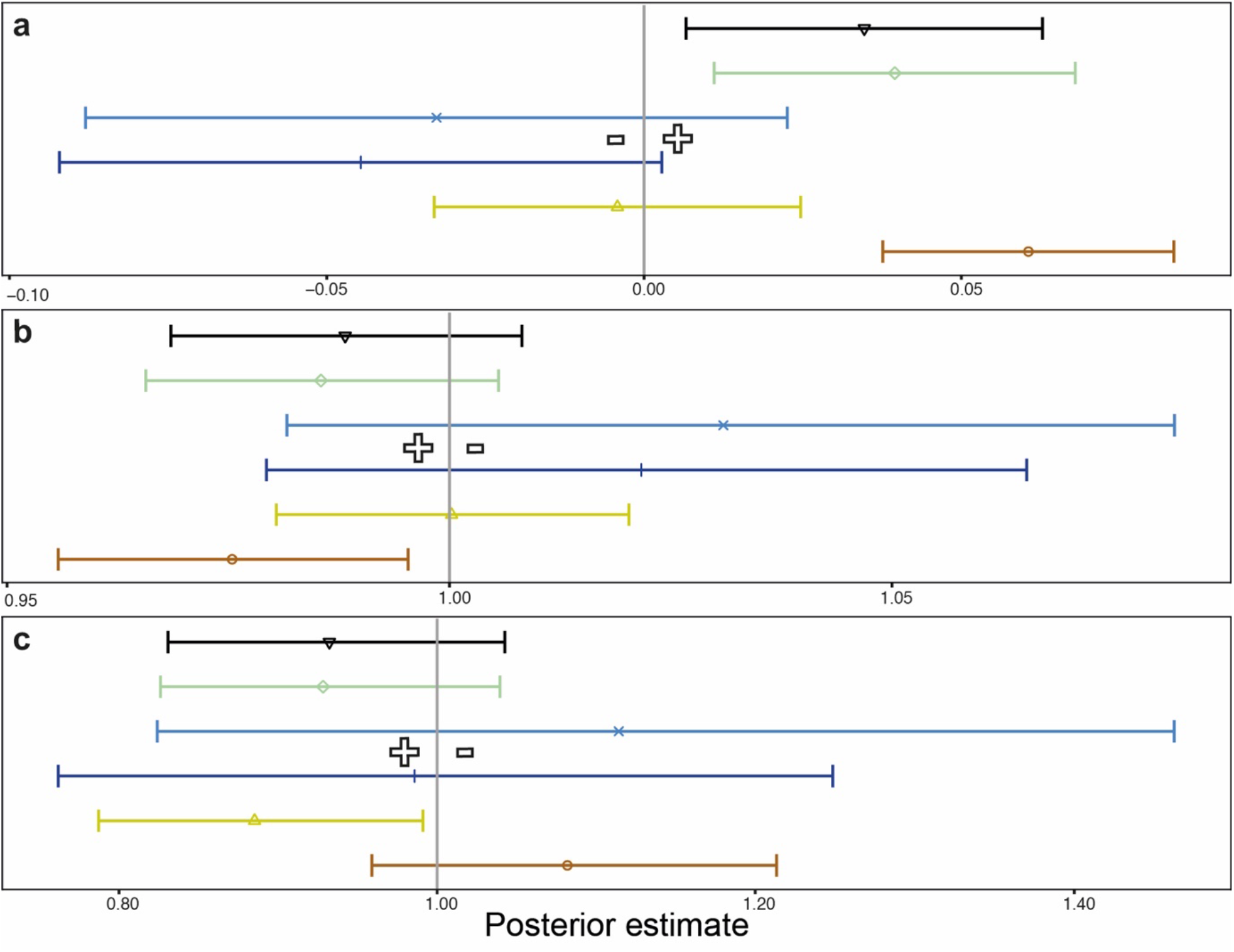
The associations between our natural habitat daily exposure rates (DER), and cognitive development, mental health and overall well-being across London. The association between (a) executive function (EF) score, (b) Strengths and Difficulties Questionnaire total difficulties score and (c) KIDSCREEN-10 Questionnaire Health-Related Quality of Life score with the DER of natural space 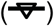, green space 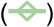, blue space level 2 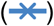, blue space level 3 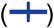, grassland 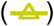 and woodland 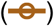. Fully adjusted model was plotted with posterior mean and 95% credible intervals (CI) and included age, area-level deprivation, ethnicity, gender, parental occupation and school type. Models with EF as the outcome were additionally adjusted for air pollution. The vertical line (grey) is the reference line and is set to zero or one depending on the model used for the outcome in analysis. Hollow plus or minus sign indicated whether the association had a positive or negative contribution to cognitive development or mental health.

To further assess the role of different types of urban natural habitat to children’s cognitive development and mental health, we characterised green space into two distinctive natural habitat types, i.e. grassland and woodland. We found that children’s cognitive development and mental health did improve with higher DER to woodland. When all other risk factors were held constant, there was a beneficial contribution to cognitive development by 0.42 (95% CI: 0.21, 0.57) points using the EF score and a reduction in the risk of emotional and behavioural problems by -0.17 (95% CI: -0.32, -0.03) points using the SDQ total difficulties score (Fig. 2 and Supplementary Figure 3). We found no improvement of overall well-being with higher DER to woodland (Fig. 2c and Supplementary Figure 3c). When comparing those children exposed to the highest level of woodland (∼38%) to those exposed to the lowest level of woodland (0%) in our study, we estimated a percent change in cognitive development of 6.83% (95% CI: 3.41, 9.11) using the EF score, and a percent change in the risk of emotional and behavioural problems of -16.36% (95% CI: -27.49, -3.50) using the SDQ total difficulties score. We found no improvement of children’s cognitive development and mental health with a higher DER to grassland with the exception of our outcome for overall well-being using the HRQoL score (Fig. 2 and Supplementary Figure 3).

### The role of other risk factors for cognition and mental health

We fitted our longitudinal models with a number of other risk factors to account for demographic, environmental and socio-economic factors that are known to influence children’s cognitive development and mental health ^5,6^. We found that our outcomes for children’s cognitive development and mental health were influenced by the child’s age, ethnic background, gender, parental occupation and type of school (Supplementary Table 2,3,4). When compared to independent schools for example, state schools were predicted to result in a negative contribution to children’s cognitive development and mental health by a percent change decrease of -5.10% (95% CI: -6.05, -4.30) using the EF score, a 10% (95% CI: 5, 15) increase in the risk of emotional and behavioural problems using the SDQ total difficulties score, and an increase in odds of exhibiting low overall well-being by 57% using the HRQoL score (95% CI: 19, 104). In addition to this, we found that air pollution influenced children’s cognitive development in most of our models (Supplementary Table 2). When removing demographic, environmental, and socio-economic factors from our models, we showed that modelled environmental variables were, in general, tenfold smaller than the contribution of our demographic and socio-economic variables (Supplementary Table 5). This stepwise exclusion of fixed effects from our models highlights the relative importance of our demographic and socio-economic variables to children’s cognitive development and mental health.

To test the robustness of our findings, we did a series of sensitivity analyses to assess which models perform best for evaluating the association between children’s cognitive development and mental health, and types of urban natural habitat. This included testing each child’s DER for (i) different buffer areas around their home and school and (ii) a different weighting based on a full day (24 hours) instead of a daytime (12 hours) weighting (See Methods). For our analyses of different buffer areas, we found that our results were consistent across different buffer areas but some models did suggest a weaker association with smaller buffer areas when compared with larger buffer areas (Supplementary Figure 1,2,3). This suggests that exposure to natural habitats further away from their home and school may play a more important role for their cognitive development and mental health than previously hypothesised by other studies^25^. When using a different weighting for our DER, we found that our models showed consistent patterns when we modelled with a DER based on a daytime or full day weighting (Supplementary Table 2,3,4).

## DISCUSSION

To our knowledge, this is the largest epidemiological study to report on the impact of natural habitat type exposure on cognitive development and mental health in children from late childhood to early adolescence. Our results showed a strong association between woodland exposure, children’s cognitive development and mental health. We also found that exposure to natural space or green space was associated with a beneficial contribution to cognitive development, while there was a weaker association for our mental health outcomes. Finally, we did not find a consistent association of blue space or grassland exposure with cognitive development and mental health.

Overall, we observed that exposure to woodland was associated with a beneficial contribution to cognitive development and a lower risk of emotional and behavioural difficulties from late childhood to early adolescence. This is in line with previous reports of positive impacts from woodland on physical and mental health^19,23,30^, with the exception of a study performed in central Scotland^31^. Forest bathing, for example, is a relaxation therapy that has been associated with physiological benefits, supporting the human immune function, reducing heart rate variability and salivary cortisol, and psychological benefits such as reduced feelings of hostility and depression^19,30^. However, the hypothetical mechanisms why we experience these psychological benefits from woodland remain unknown. Higher audio-visual exposure through vegetation and animal abundance has been documented to improve mental health, of which both features are expected in higher abundance in woodland^17,32^. Even though our results show that urban woodland is associated with children’s cognitive development and mental health, the mechanistic pathway to explain this association in urban areas remains unknown.

Our results also showed that exposure to natural space or green space was associated with a beneficial contribution to children’s cognitive development, which was consistent with previous studies^12,33^. For the mental health outcomes, our findings for weaker associations with exposure to natural space or green space is consistent with the variability in these relationships found in previous studies^10,11,21,34^. One reason explaining this variability may be that most studies, including this study, do not account for the quality of green space, which has been proposed as more important than the quantity of green space^35^. Nevertheless, systematic reviews suggest that nature positively influences mental health; even though, evidence is often limited to cross-sectional studies, and inadequate particularly for children^28^.

We did not find a consistent association between blue space exposure, cognitive development and mental health. However, we cannot dismiss that blue space may be associated with cognitive development and mental health as other studies have found associations^25,36^. In our study of 3,568 children, 66.8% had no blue space within a 250 m buffer area, showing that the amount of blue space surrounding children’s residence and school was low regardless. One explanation for this weak association may be the difference of natural habitat composition from one city to the other, potentially changing people’s relationship with nature. Residents in coastal cities, for example, are assumed to have a different relationship with blue space compared to cities inland where blue space may be less abundant. Alternatively, inconsistencies may be the result of different sampling techniques. For example, other studies have used self-reported blue space visitation rates or blue space visibility and found associations with behavioural difficulties and psychological distress^25,36^. Inconsistencies because of different sampling techniques make it difficult to harmonize results into a consistent framework, but to date there has been no comprehensive analysis allowing for harmonisation in green space mental health research.

Urban nature may have multiple benefits for human health and well-being, including changing air pollution, physical and mental health, and social well-being. Optimisation of nature by decision-makers to maximise cognitive development and mental health benefits from nature exposure may have synergies or trade-offs with other ES, especially in urban areas where space is limited. For example, our results found that higher levels of woodland were associated with enhanced cognitive development and mental health, which can be synergistic with climate mitigation efforts to sequester carbon dioxide^37^, but can have trade-offs with children’s physical health when woodland were to replace open spaces specifically designated for physical activity^38^. The lack of a conceptual framework between nature, cognitive development and mental health is also a key hindrance for the inclusion of these ES in international standardised environmental frameworks such as the System of Environmental-Economic Accounting Experimental Ecosystem Accounting, nor in the revision which is currently taking place^39^. The complexity of interactions and lack of knowledge on ES related to cognitive and mental benefits has made it difficult to identify which decisions should be made regarding management of urban nature, a critical knowledge gap given the increase in the global urban population this century^1^.

The study has several strengths. It used a high-quality cohort dataset of considerable size for children during late childhood and early adolescence, a subset of the urban population which is often understudied. This large sample had substantial spatio-temporal diversity on an urban scale for the Greater London metropolitan area with sufficient statistical power to investigate interactions. The study used clinically validated instruments to define children’s cognitive development and mental health. It also used geographical information of very high resolution to develop metrics of natural habitat DER such as NDVI at 10 m resolution and LiDAR at 2 m resolution. Previous studies have used conventional satellite remote-sensing data for establishing associations between green space, cognitive development and mental health. In this study however, we developed an objective measure of exposure to natural habitat types as a proxy for characterising different natural habitats. This study also adjusted for other potential confounders through objective measures of exposure to air pollution, socioeconomic status and other individual-level confounders.

Despite of our large sample size using a rigorous longitudinal study design, our results could be influenced by a number of potentially confounding factors. For example, we cannot necessarily assume that children’s DER to natural space leads to increased use of natural space, as the quality of natural space could also play a role^25,35^. Our data also did not provide information on when exactly children moved to a new residence between baseline and follow-up, which may influence our measure of DER. Additionally, although we adjusted for individual and area-level socioeconomic status through parental occupation and the Carstairs deprivation index, unmeasured risk factors such as higher crime rates, housing prices, or fewer social advantages may also influence our results^40^. Finally, although our study importantly sheds light on the role of natural habitat types for cognitive development and mental health, it also highlights the gap in understanding the mechanisms of the benefits of woodland over other types of natural habitat.

## CONCLUSION

Our study showed that higher levels of woodland were associated with a beneficial contribution to cognitive development and a lower risk of emotional and behavioural problems from late childhood to early adolescence. These findings contribute to our understanding of urban natural habitats as an important risk factor for children’s cognitive development and mental health. Ensuring fair and equitable access to woodland could be an important tool to manage and minimise cognitive development and mental health problems, especially in children who are in the midst of their development into adulthood. Lower access to woodland may also be an added risk factor among more vulnerable groups in society. Our findings contribute to our understanding of the physical and monetary valuation of cognitive and mental health benefits received from urban nature, suggesting that not every type of natural space may contribute equally to cognitive development and mental health benefits. As part of the growing human health green space research, our study concludes that understanding people’s local relationship with nature may be a key component to understand its association with cognitive development and mental health. This should be considered as part of ongoing efforts to sustainably develop urban nature and to standardise international measurement and environmental accounting frameworks for ES connected to cognitive development and mental health benefits.

## METHODS

### Study population

We use data from SCAMP^41^, a longitudinal cohort study established to investigate how the cognitive development and behaviour of children across the London metropolitan area during late childhood and early adolescence might be affected by use of mobile phones and other technologies that use radio waves. A baseline and follow-up school visit were carried out between 2014 and 2018 with a time gap of approximately 2 years between the baseline and follow-up visit for each school. Initially, 6,612 children participated to the baseline assessment, and 5,208 children participated to the follow-up. A total of 3,791 children participated to both the baseline and follow-up assessment, and for our analysis we used a subset of 3,568 children who had a known home address during the baseline and follow-up assessment (Fig. 1a). This subset excluded 8 schools due to low sampling size (< 15 children per school). Included children were on average 12 and 14.2 years old during the baseline and follow-up assessment respectively, and 57.9% of them were female. The children (*n* = 3,568) were part of 31 schools across London, of which 12 were independent schools and 19 were state schools. Of the 31 participating schools, 3 were located outside the Greater London Authority (GLA) administrative area (Fig. 1a). During the assessments, information was gathered on age, gender (two levels: female or male), ethnicity (five levels: White, Black, Asian, mixed or other), school type (two levels: state or independent), parental occupation (five levels: managerial/professional occupations, intermediate occupations, small employers/own account workers, lower supervisory/technical occupations or semi-routine/routine occupations)^42^, and area-level deprivation (divided in quintiles ranging from category 1 ‘least deprived’ to category 5 ‘most deprived’). Area-level deprivation was based on the Carstairs deprivation index to identify socioeconomic confounding^43^. The scores were standardised to the area in which the child lived and not to the child itself in order to reflect the material deprivation of the area in relation to neighbouring areas. It is measured based on four variables from the United kingdom (UK) Office of National Statistics 2011 Census: proportion of low social class, lack of car ownership, household overcrowding and male unemployment^44^. Further characteristics of the study population are presented in Supplementary Table 6. All parents or guardians signed the informed consent and the study was approved (REC reference: 14/NW/0347) by the Health Research Authority NKES Committee North West - Haydock. Study population data are not publicly available for data protection issues. To request access to the data, contact M. B. Toledano at.

### Outcomes

Children’s cognitive development was assessed through the EF, a composite score of three computerised tests (i.e. Backward Digit Span [BDS], Spatial Working Memory [SWM] and Trail Making Task [TMT])^45–47^. EF composite was only calculated for children who completed all three contributing tasks. We derived the EF composite at baseline by taking an average of Z-scores for the key performance measure for each EF task^48^. The composite score at follow-up was derived by taking an average of scores for the same EF tasks, equivalently adjusted by the mean and SD from baseline performance. Z-scores and adjusted values were calculated across the whole population at each time point. TMT and SWM values were reverse coded prior to taking the average. EF values were continuous and higher EF values indicated better cognitive performance (Fig. 1b).

We assessed children’s mental health from the self-reported SDQ and the KIDSCREEN-10 Questionnaire taken by each child^49^. The SDQ total difficulties score assesses the emotion and behaviour of children and was calculated by summing the scores for the four difficulties subscales on emotional problems, conduct, hyperactivity and peer problems. Each subscale comprised of five items that can be scored 0, 1 or 2 and each subscale score can therefore range from 0 to 10. An SDQ total difficulties score was treated as count data where a higher value represented more behavioural difficulties (Fig. 1c)^49^.

The KIDSCREEN-10 HRQoL score consists of 10 self-reported items covering physical, psychological and social dimensions of well-being, with children indicating the frequency or severity of each item on a 5 point Likert scale (1 = never/not at all, 2 = almost never/slightly, 3 = sometimes/moderately, 4 = almost always/very and 5 = always/extremely). Totals of these 10 items were summed with higher values indicating better HRQoL. Rasch person parameters were assigned to each possible total based on the Rasch model, a psychometric model commonly used for measurements of categorical data^50^. The Rasch-scaled single score of HRQoL was then transformed into scores with a mean of 50 and a standard deviation of approximately 10, where a higher score indicates a better HRQoL (Fig. 1d)^50^. In line with previous studies, binary cut-offs were applied based on the lower 10% of the sample distribution (i.e. baseline and follow-up mean below 39.28 and 36.51, respectively) to identify children with noticeably low overall well-being (two levels: 0 - high overall well-being and 1 - low overall well-being)^51^. All data on children’s cognitive development, mental health and overall well-being were gathered using Psytools software (Delosis Ltd., London).

### Quantification of natural habitat composition

Our exposure assessment of natural habitat was based on a three-tier stepwise characterisation: (1) natural space, (2) green and blue space, and (3) grassland and woodland. We used different data sources to quantify the natural habitats surrounding the residential and school area of each child. Firstly, we generated a NDVI spatial layer of our study area using data from the Sentinel-2 satellite at 10 m spatial resolution^52^. NDVI is a unit-less index of relative overall vegetation density and quality based on differential surface reflectance in the red and near infrared regions with low values indicating sparse vegetation, and high values indicating dense vegetation^53^. We generated our NDVI map by using Google Earth Engine to filter out satellite data between July 1^st^ 2015 and July 1^st^ 2017 for images with less severe cloud cover (<5%)^54^. Images covering the same area at different dates were then mosaicked into a single complete and cloud-free image of NDVI (Supplementary Figure 4a). Secondly, we created a spatial layer from surface and tidal water maps from the Ordnance Survey (OS) Open Map, a large-scale digital map covering Great Britain (Supplementary Figure 4b)^55^.

To further assess fine-scale habitat composition within green space, we used Light Detection and Ranging (LiDAR) data^56^ from the Environment Agency (data.gov.uk, accessed July 2^nd^ 2018, licensed under the Open Government Licence 3.0) (Supplementary Figure 4c). We used the LiDAR Composite Digital Surface Model and Digital Terrain Model at 2 m spatial resolution to estimate object height across our study area. Within green space, we split vegetation in two height strata: 0 - 1 m and (>1 m) and we assumed that vegetation between 0 - 1 m was predominantly grassland, and >1 m was woodland^56^.

We calculated each child’s proportionate DER to each natural habitat characterisation in buffer areas of 50 m, 100 m, 250 m and 500 m around the residential and school area:

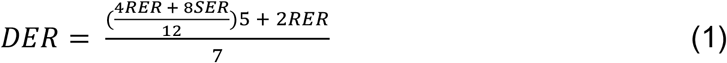

where DER is the daily exposure rate, RER is the residential exposure rate and SER is the school exposure rate. We assumed each child spent the weekend in their residential area, while we weighted weekdays by the daytime (12 hours) children were assumed to spend at home (4 hours) and at school (8 hours). We selected different buffer areas to assess the consistency of our results in a comparable manner with previous studies^11,12,25^. Based on the above formula, we calculated natural space DER by combining our NVDI and water maps. Then, we calculated green and blue space DER by using our NDVI and water maps separately. Finally, we calculated grassland and woodland DER by combining our NDVI and height strata map. The different spatial resolutions of our NDVI and height strata map resulted in classification errors where pixels were misclassified as grassland or woodland when in fact it was part of the built environment. To correct for this, we excluded buildings from these maps using the buildings feature from OS Open Map (Supplementary Figure 4d)^55^. It was difficult to use blue space DER of the 3,568 participants because 2,383 children (66.8%) had, for example, no blue space within 250 m. We therefore reclassified blue space into tertiles (three levels: level 1 - no blue space, level 2 - blue space with a DER below the mean, and level 3 - blue space with a DER above the mean).

### Quantification of outdoor air pollution

Considering the ability of nature to mitigate local air pollution^57^, we hypothesised that exposure to air pollution could be an underlying confounder between nature exposure and cognitive development^58^. We based our exposure assessment of air pollution on emission estimates of key air pollutants using the London Atmospheric Emission Inventory (LAEI) 2016 from GLA and Transport for London (data.london.gov.uk, accessed February 27^th^ 2020, licensed under the UK Open Government Licence 2.0). The LAEI estimated ground level concentrations of four air pollutants (nitrogen dioxide [NO_2_], nitrogen oxides [NO_x_] and particulate matter with a diameter of 10 microns [PM_10_] or 2.5 microns or less [PM_2.5_]) using an atmospheric dispersion model, and covered Greater London, as well as areas outside Greater London up to the M25 motorway. A total of 3,305 children (out of 3,568 children) were located within the M25 motorway and therefore eligible to measure ambient air pollution. Similar to the quantification of natural habitat composition, we calculated each child’s average DER to each air pollutant in buffer areas of 50 m, 100 m, 250 m and 500 m around the residential and school area following equation 1. The Pearson’s correlation coefficient among DERs ranged from 0.95 (between NO_2_ and PM_10_) to 0.98 (between NO_2_ and NO_x_) (Supplementary Table 7). To avoid multicollinearity, we used NO_2_ DER as it is a commonly used proxy for traffic-related air pollution.

### Statistical analyses

Our modelling framework consisted of Bayesian longitudinal regression models to examine the relationship between natural habitat DERs, and our cognitive development and mental health outcomes. Inference was performed using Integrated Nested Laplace Approximation (INLA)^59^. The Pearson’s correlation coefficient among natural habitat DERs ranged from 0.38 (between grassland and woodland) to 0.99 (between natural space and green space) (Supplementary Table 1). The high Pearson’s correlation coefficient was not considered a problem because we performed separated analyses for the different DERs. In particular, we developed three multilevel modelling structures including these as fixed effects, where Model I (M I) included natural space DER, Model II (M II) included green and blue space DER, and Model III (M III) included grassland and woodland DER. Our outcomes consisted of two repeated measures per child, i.e. a baseline and follow-up measure. We assumed a Gaussian, Poisson and Binomial distribution for the EF score, SDQ total difficulties score and HRQoL score, respectively. We included a random effect term for child identifier to allow for between-child variance, while we used a random effect term for tests at the time of visit (two levels: baseline or follow-up) for each child to introduce correlation among the repeated measurements. School was not added as an additional random effect in our multilevel model because it did not improve the model fit, and three different cross-validation techniques were used for model comparison and selection (Supplementary Table 8,9,10). Fully adjusted models included natural habitat DERs, age, area-level deprivation, ethnicity, gender, parental occupation and school type, and models with EF score were additionally adjusted for air pollution. Additionally, we did a stratified analysis to investigate potential changes in point estimates and avoid potential bias from over adjustment (four levels: unadjusted, adjusted for ethnicity and school type, adjusted for socioeconomic factors and adjusted for all risk factors) (Supplementary Figure 1,2,3). A detailed description of the model structures is given in Supplementary Methods 1. Prior to the longitudinal analysis, a cross-sectional analysis of the baseline cohort was done which was qualitatively similar to the longitudinal results and is therefore not further discussed (Supplementary Methods 2 and Supplementary Figure 5).

We performed the following sensitivity analyses to determine the best models for evaluating the association with natural habitat DER by fitting additional Bayesian mixed-effect models for (i) the association with different buffer areas (Supplementary Figure 1,2,3) and (ii) the association with a different weighting of natural habitat DERs based on a full day (24 hours) instead of a daytime (12 hours) weighting where we assumed children spend 16 hours at home and 8 hours at school during the weekdays (Supplementary Table 2,3,4). In the main text, unless stated otherwise, results were based on fully adjusted models with natural habitat DERs with a daytime weighting and measured in buffer areas of 250 m as we found no strong difference when measuring at different buffer areas, and between daytime or full day weighting. We did all data processing and statistics in Python 3.7.3., ArcGIS 10.7 and R 4.0.0 via RStudio using the packages brinla, ggplot2, ggpubr, R-INLA, MBA, raster, rgdal, sp and spdep^60^.

## Data Availability

Study population and environmental exposure data around each child's home and school are not publicly available for data protection issues. To request access to the data, contact M. B. Toledano at. Environmental data that are the basis for our environmental exposure data are available at  github.com/MikaelMaes/HumanExposure.git. This environmental data are based on publicly available sources. Sentinel-2 satellite data are available using Google Earth Engine at earthengine.google.com. Buildings, surface water and tidal water data from the OS Open Map are available at ordnancesurvey.co.uk. LiDAR data from the Environment Agency are available at data.gov.uk. Air pollution estimates using the London Atmospheric Emissions Inventory 2016 from the Greater London Authority and Transport for London are available at data.london.gov.uk. The full model outputs that support the findings of this study are available in the Supplementary Information.

https://github.com/MikaelMaes/HumanExposure

## DATA AVAILABILITY

Study population and environmental exposure data around each child’s home and school are not publicly available for data protection issues. To request access to the data, contact M. B. Toledano at. Environmental data that are the basis for our environmental exposure data are available at github.com/MikaelMaes/HumanExposure.git. This environmental data are based on publicly available sources. Sentinel-2 satellite data are available using Google Earth Engine at earthengine.google.com. Buildings, surface water and tidal water data

from the OS Open Map are available at ordnancesurvey.co.uk. LiDAR data from the Environment Agency are available at data.gov.uk. Air pollution estimates using the LAEI 2016 from GLA and Transport for London are available at data.london.gov.uk. The full model outputs that support the findings of this study are available in the Supplementary Information.

## CODE AVAILABILITY

The source code to compute NDVI from satellite data using Google Earth Engine is available at earthengine.google.com. The code for processing raw LiDAR data, creating our environmental exposure variables and modelling our data is available at github.com/MikaelMaes/HumanExposure.git.

## ACKNOWLEDGEMENTS

We thank Professor Marta Blangiardo (Imperial College London) and Dr Rory Gibb (University College London) for feedback on the statistical analyses. This study is supported by funding of the London Natural Environment Research Council Doctoral Training Program (NE/L002485/1), the MRC Centre for Environment and Health (MR/L01341X/1) based at Imperial College London and the NIHR Health Protection Research Unit in the Health Impact of Environmental Hazards, based at King’s College London and Imperial College London, in partnership with Public Health England (PHE) (HPRU-2012-10141). SCAMP is independent research funded by the National Institute for Health Research (NIHR) Policy Research Program (PRP) (Secondary School Cohort Study of Mobile Phone Use and Neurocognitive and Behavioural Outcomes/091/0212) via the Research Initiative on Health and Mobile Telecommunications, a partnership between public funders and the mobile phone industry. An extension to SCAMP is funded by NIHR PRP. The funders of the study had no role in the design or conduct of the study nor the reporting of the SCAMP study results. M.B.T. chair and the work in this paper is supported in part by a donation from Marit Mohn to Imperial College London to support Population Child Health. The views expressed in this paper are those of the authors and not necessarily those of the NIHR, DHSC, PHE or any other funder.

## AUTHOR CONTRIBUTIONS

M.J.A.M., K.E.J. and M.T.B. conceived the study and analysed the results. E.B. provided data on cognitive development. M.J.A.M. coded the models, performed the simulations and wrote the manuscript with substantial contributions from all the authors.

## COMPETING INTERESTS

The authors declare no competing interests.

## Notes

### Competing Interest Statement

The authors have declared no competing interest.

### Author Declarations

All parents or guardians signed the informed consent and the study was approved (REC reference: 14/NW/0347) by the Health Research Authority NKES Committee North West - Haydock.

## REFERENCES

1. UN DESA. World Urbanization Prospects: The 2018 Revision (ST/ESA/SER.A/420). (2019).

2. Giles-Corti, B. et al. City planning and population health: a global challenge. Lancet 388, 2912–2924 (2016).

3. Okkels, N., Kristiansen, C. B., Munk-Jørgensen, P. & Sartorius, N. Urban mental health. Curr. Opin. Psychiatry 31, 258–264 (2018).

4. Robbins, R. N., Scott, T., Joska, J. A. & Gouse, H. Impact of urbanization on cognitive disorders. Curr. Opin. Psychiatry 32, 210–217 (2019).

5. Afifi, M. Gender differences in mental health. Singapore Med. J. 48, 385–391 (2007).

6. Guhn, M., Emerson, S. D., Mahdaviani, D. & Gadermann, A. M. Associations of Birth Factors and Socio-Economic Status with Indicators of Early Emotional Development and Mental Health in Childhood: A Population-Based Linkage Study. Child Psychiatry Hum. Dev. 51, 80–93 (2020).

7. Torales, J., O’Higgins, M., Castaldelli-Maia, J. M. & Ventriglio, A. The outbreak of COVID-19 coronavirus and its impact on global mental health. Int. J. Soc. Psychiatry 66, 317–320 (2020).

8. Holmes, E. A. et al. Multidisciplinary research priorities for the COVID-19 pandemic: a call for action for mental health science. The Lancet Psychiatry vol. 7 547–560 (2020).

9. GLA. London Mental Health: The invisible costs of mental ill health. https://www.london.gov.uk/sites/default/files/gla_migrate_files_destination/Mentalhealthreport.pdf (2014).

10. Sarkar, C., Webster, C. & Gallacher, J. Residential greenness and prevalence of major depressive disorders: a cross-sectional, observational, associational study of 94?879 adult UK Biobank participants. Lancet. Planet. Heal. 2, e162– e173 (2018).

11. Engemann, K. et al. Residential green space in childhood is associated with lower risk of psychiatric disorders from adolescence into adulthood. Proc. Natl. Acad. Sci. 201807504 (2019) doi:10.1073/PNAS.1807504116.

12. Dadvand, P. et al. Green spaces and cognitive development in primary schoolchildren. Proc. Natl. Acad. Sci. U. S. A. 112, 7937–42 (2015).

13. Haines-Young, R. & Potschin, M. Common International Classification of Ecosystem Services (CICES) V5.1. (2018).

14. Franco, L. S., Shanahan, D. F. & Fuller, R. A. A Review of the Benefits of Nature Experiences: More Than Meets the Eye. Int. J. Environ. Res. Public Health 14, 864 (2017).

15. Cox, D. T. C. et al. Skewed contributions of individual trees to indirect nature experiences. Landsc. Urban Plan. 185, 28–34 (2019).

16. Ulrich, R. View Through a Window May Influence Recovery from Surgery. Science (80-.). 224, 224–225 (1984).

17. Irvine, K. N. et al. Green space, soundscape and urban sustainability: an interdisciplinary, empirical study. Local Environ. 14, 155–172 (2009).

18. Weber, S. T. & Heuberger, E. The Impact of Natural Odors on Affective States in Humans. Chem. Senses 33, 441–447 (2008).

19. Li, Q. Effect of forest bathing trips on human immune function. Environ. Health Prev. Med. 15, 9–17 (2010).

20. Rook, G. A., Raison, C. L. & Lowry, C. A. Can we vaccinate against depression? Drug Discov. Today 17, 451–458 (2012).

21. Markevych, I. et al. Access to urban green spaces and behavioural problems in children: Results from the GINIplus and LISAplus studies. Environ. Int. 71, 29–35 (2014).

22. Taylor, M. S., Wheeler, B. W., White, M. P., Economou, T. & Osborne, N. J. Research note: Urban street tree density and antidepressant prescription rates—A cross-sectional study in London, UK. Landscape and Urban Planning vol. 136 (2015).

23. Akpinar, A., Barbosa-Leiker, C. & Brooks, K. R. Does green space matter? Exploring relationships between green space type and health indicators. Urban For. Urban Green. 20, 407–418 (2016).

24. Cox, D. T. C., Shanahan, D. F., Hudson, H. L., Fuller, R. A. & Gaston, K. J. The impact of urbanisation on nature dose and the implications for human health. Landsc. Urban Plan. 179, 72–80 (2018).

25. Amoly, E. et al. Green and Blue Spaces and Behavioral Development in Barcelona Schoolchildren: The BREATHE Project. Environ. Health Perspect. 122, 1351–1358 (2014).

26. Astell-Burt, T. & Feng, X. Association of Urban Green Space With Mental Health and General Health Among Adults in Australia. JAMA Netw. Open 2, e198209 (2019).

27. Barton, J. & Pretty, J. What is the Best Dose of Nature and Green Exercise for Improving Mental Health?? A Multi -Study Analysis. Environ. Sci. Technol. 44, 3947–3955 (2010).

28. Gascon, M. et al. Mental health benefits of long-term exposure to residential green and blue spaces: A systematic review. Int. J. Environ. Res. Public Health 12, 4354–4379 (2015).

29. PHE. The mental health of children and young people in London. (2016).

30. Morita, E. et al. Psychological effects of forest environments on healthy adults: Shinrin-yoku (forest-air bathing, walking) as a possible method of stress reduction. Public Health 121, 54–63 (2007).

31. Thompson, C. W. Woodland improvements in deprived urban communities: how does this build resilience? Eur. J. Public Health 27, (2017).

32. Hedblom, M., Heyman, E., Antonsson, H. & Gunnarsson, B. Bird song diversity influences young people’s appreciation of urban landscapes. Urban For. Urban Green. 13, 469–474 (2014).

33. Liao, J. et al. Residential exposure to green space and early childhood neurodevelopment. Environ. Int. 128, 70–76 (2019).

34. Picavet, H. S. J. et al. Greener living environment healthier people? Exploring green space, physical activity and health in the Doetinchem Cohort Study. Prev. Med. (Baltim). 89, 7–14 (2016).

35. Francis, J., Wood, L. J., Knuiman, M. & Giles-Corti, B. Quality or quantity? Exploring the relationship between Public Open Space attributes and mental health in Perth, Western Australia. Soc. Sci. Med. 74, 1570–1577 (2012).

36. Nutsford, D., Pearson, A. L., Kingham, S. & Reitsma, F. Residential exposure to visible blue space (but not green space) associated with lower psychological distress in a capital city. Health Place 39, 70–78 (2016).

37. McPherson, E. G., Xiao, Q. & Aguaron, E. A new approach to quantify and map carbon stored, sequestered and emissions avoided by urban forests. Landsc. Urban Plan. 120, 70–84 (2013).

38. Lachowycz, K., Jones, A. P., Page, A. S., Wheeler, B. W. & Cooper, A. R. What can global positioning systems tell us about the contribution of different types of urban greenspace to children’s physical activity? Health Place 18, 586–94 (2012).

39. UN. System of Environmental-Economic Accounting - Ecosystem Accounting: Draft for the Global Consultation on the complete document. (2020).

40. Tarling, R. & Roger, R. D. Socio-Economic Determinants of Crime Rates: Modelling Local Area Police-Recorded Crime. Howard J. 55, 207–225 (2016).

41. Toledano, M. B. et al. Cohort Profile: The Study of Cognition, Adolescents and Mobile Phones (SCAMP). Int. J. Epidemiol. 48, 25–26l (2018).

42. Rose, D., Pevalin, D. J. & O’Reilly, K. The National Statistics Socio-economic Classification: Origins, Development and Use. https://www.researchgate.net/publication/312200977 (2005).

43. Carstairs, V. & Morris, R. Deprivation and health in Scotland. Health Bull. (Raleigh). 48, 162–75 (1990).

44. Office of National Statistics. 2011 Census aggregate data. https://www.ons.gov.uk/census/2011census (2012).

45. Luciana, M. & Nelson, C. A. Assessment of neuropsychological function through use of the Cambridge Neuropsychological Testing Automated Battery: Performance in 4-to 12-year-old children. Dev. Neuropsychol. 22, 595–624 (2002).

46. Dumontheil, I. & Klingberg, T. Brain Activity during a Visuospatial Working Memory Task Predicts Arithmetical Performance 2 Years Later. Cereb. Cortex 22, 1078–1085 (2012).

47. Tombaugh, T. N. Trail Making Test A and B: Normative data stratified by age and education. Arch. Clin. Neuropsychol. 19, 203–214 (2004).

48. Burgess, P. W. Theory and methodology in executive function research. in Methodology of Frontal and Executive Function 79–113 (Taylor and Francis, 2004). doi:10.4324/9780203344187-8.

49. Goodman, R., Meltzer, H. & Bailey, V. The Strengths and Difficulties Questionnaire: a pilot study on the validity of the self-report version. Int. Rev. Psychiatry 15, 173–177 (2003).

50. The KIDSCREEN Group Europe. The Kidscreen questionnaires––quality of life questionnaires for children and adolescents. (Pabst Science Publishers, 2006).

51. Berman, A. H., Liu, B., Ullman, S., Jadbäck, I. & Engström, K. Children’s Quality of Life Based on the KIDSCREEN-27: Child Self-Report, Parent Ratings and Child-Parent Agreement in a Swedish Random Population Sample. PLoS One 11, e0150545 (2016).

52. ESA. Sentinel-2 User Handbook. (2015).

53. Gascon, M. et al. Normalized difference vegetation index (NDVI) as a marker of surrounding greenness in epidemiological studies: The case of Barcelona city. Urban For. Urban Green. 19, 88–94 (2016).

54. Gorelick, N. et al. Google Earth Engine: Planetary-scale geospatial analysis for everyone. Remote Sens. Environ. 202, 18–27 (2017).

55. OS. Open Map - Local. http://os.uk (2019).

56. Miura, N. & Jones, S. D. Characterizing forest ecological structure using pulse types and heights of airborne laser scanning. Remote Sens. Environ. 114, 1069–1076 (2010).

57. Dadvand, P. et al. The association between greenness and traffic-related air pollution at schools. Sci. Total Environ. 523, 59–63 (2015).

58. Sunyer, J. et al. Association between Traffic-Related Air Pollution in Schools and Cognitive Development in Primary School Children: A Prospective Cohort Study. PLOS Med. 12, e1001792 (2015).

59. Rue, H., Martino, S. & Chopin, N. Approximate Bayesian inference for latent Gaussian models by using integrated nested Laplace approximations. J. R. Stat. Soc. Ser. B (Statistical Methodol. 71, 319–392 (2009).

60. RStudio Team. RStudio: Integrated Development for R. RStudio. (2015).

