## Supplementary information for "Benefits of natural habitat particularly woodland on children’s cognition and mental health"

### Supplementary information: Benefits of natural habitat particularly woodland on children's cognitive development and mental health

Mikaël J. A. Maes[1,2,3,4], Monica Pirani[3], Elizabeth R. Booth[5], Chen Shen[3], Ben Milligan[6,7], Kate E. Jones[2] and Mireille B. Toledano[3,4]

[1]Department of Geography, University College London, Pearson Building, Gower Street, London, WC1E 6BT, United Kingdom.
[2]Centre for Biodiversity and Environment Research, Department of Genetics, Evolution and Environment, University College London, Gower Street, London, WC1E 6BT, United Kingdom.
[3]MRC Centre for Environment and Health, School of Public Health, Faculty of Medicine, Imperial College London, Norfolk Place, London W2 1PG, United Kingdom
[4]Mohn Centre for Children's Health and Wellbeing, School of Public Health, Faculty of Medicine, Imperial College London, Norfolk Place, London W2 1PG, United Kingdom.
[5]Centre for Educational Neuroscience, Department of Psychological Sciences, Birkbeck College, Malet Street, London, WC1E 7HX, United Kingdom
[6]Institute for Sustainable Resources, University College London, Central House, 14 Upper Woburn Place, London, WC1H 0NN, United Kingdom
[7]University of New South Wales Law School, Law Building, UNSW Sydney, NSW 2052, Australia

| Supplementary information – Table of Contents | Page(s) |
|---|---|
| **Supplementary Methods 1**<br>• Detailed description of the model structures for our longitudinal analysis.<br>**Supplementary Methods 2**<br>• Detailed description of the model structures for our cross-sectional analysis | 3-6 |
| **Supplementary Figures**<br>• **Supplementary Figure 1.** Comparison of different buffer areas to investigate the association between natural space daily exposure rates (DER) and cognitive development and mental health from late childhood to early adolescence.<br>• **Supplementary Figure 2.** Comparison of different buffer areas to investigate the association between green and blue space DER and cognitive development and mental health from late childhood to early adolescence.<br>• **Supplementary Figure 3.** Comparison of different buffer areas to investigate the association between grassland and woodland DER and cognitive development and mental health from late childhood to early adolescence.<br>• **Supplementary Figure 4.** Environmental datasets used to quantify natural habitat exposure.<br>• **Supplementary Figure 5.** The associations between our environmental DER, and cognitive performance and mental health across London. | 7-11 |
| **Supplementary Tables**<br>• **Supplementary Table 1.** Median (Q1, Q3) and Pearson's correlation coefficient between estimates of natural habitat DER.<br>• **Supplementary Table 2.** Comparison of fully adjusted models with the executive function (EF) and environmental DER based on daytime (12 hrs) or full day (24 hrs) weighting.<br>• **Supplementary Table 3.** Comparison of fully adjusted models with Strengths and Difficulties Questionnaire (SDQ) total difficulties score and environmental DER based on daytime (12 hrs) or full day (24 hrs) weighting.<br>• **Supplementary Table 4.** Comparison of fully adjusted models with KIDSCREEN-10 Questionnaire Health-Related Quality of Life (HRQoL) score and environmental DER based on daytime (12 hrs) or full day (24 hrs) weighting.<br>• **Supplementary Table 5.** Contribution of risk factor groups based on the difference in pseudo R-squared between the full fixed-effects only Model I (M I) and M I excluding environmental, demographic or socioeconomic variables.<br>• **Supplementary Table 6.** Characteristics of the baseline and follow-up cohort.<br>• **Supplementary Table 7.** Median (Q1, Q3) and Pearson's correlation coefficient between estimates of air pollution DER.<br>• **Supplementary Table 8.** Cross validation results testing different models for the EF.<br>• **Supplementary Table 9.** Cross validation results testing different models for the SDQ total difficulties score.<br>• **Supplementary Table 10.** Cross validation results testing different models for the KIDSCREEN-10 Questionnaire HRQoL score. | 12-24 |

#### Supplementary Methods 1

##### Executive function (EF)

We treated the EF as a continuous variable and we therefore modelled the outcome EF with a Gaussian distribution. We started by specifying $\mathrm{Y}_{\mathrm{ij}}^{\mathrm{EF}}$ as the measured composite score of three cognitive tests (i.e. Backward Digit Span, Spatial Working Memory and Trail Making Task) measured during baseline and follow-up assessments. As the EF was characterized by tests with different scales, we z-standardized the tests to make them comparable:

$$z_i = \frac{x_i - \mu}{\sigma} \quad (1)$$

As $Y_{ij}^{EF}$ was a continuous variable and can assume any value after standardization, it was reasonable to assume a Gaussian distribution with $j = 1, 2$ (time of baseline and follow-up assessment) and $i = 1, \dots, I = 3{,}568$ (total number of children in this study):

$$Y_{ij}^{EF} \sim N(\mu_{ij}^{EF},\ \sigma_{EF}^2) \quad (2)$$

where $\sigma_{\mathrm{EF}}^2$ was the variance. On $\mu_{ij}^{EF}$, we specified a linear model:

$$\mu_{ij}^{EF} = \beta_0 + \eta_{ij} + \beta_1 X_{ij}^{age} + \beta_2 X_{ij}^{air} + \beta_3 X_{ij}^{area} + \beta_4 X_i^{ethn} + \beta_5 X_i^{gender} + \beta_6 X_{ij}^{nattyp} + \beta_7 X_i^{par} + \beta_8 X_i^{schtyp} + \varepsilon_i \quad (3)$$

where $\beta_0$ was the global intercept, $\beta_1, \dots, \beta_8$ were the regression coefficients associated with the covariates, $\varepsilon_i$ was the random effect for child $i$ and $\eta_{ij}$ was the random effect for time $j$ nested in child $i$. Air = air pollution; area = area-level deprivation; ethn = ethnicity; nattyp = natural habitat type; par = parental occupation; schtyp = school type.

##### Strengths and Difficulties Questionnaire (SDQ) total difficulties score

We treated SDQ total difficulties score as count data and we therefore modeled the outcome SDQ total difficulties score with a Poisson distribution.

We started by specifying $Y_{ij}^{TDS}$ as the observed number of behavioral difficulties with $j = 1, 2$ (time of baseline and follow-up assessment) and $i = 1, \dots, I =$ 3,568 (total number of children in this study) and specified the Poisson model:

$$Y_{ij}^{TDS} \sim Poisson\ (\lambda_{ij}^{TDS}\ E_{ij}^{TDS}) \tag{4}$$

where $E_{ij}^{TDS}$ represented the expected number of behavioral difficulties (included in the model as an offset in the log scale) and $\lambda_{ij}^{TDS}$ represented the log relative risk of behavioral difficulties. We therefore specified a regression model on the log link transformed $\lambda_{ij}^{TDS}$:

$$log\ (\lambda_{ij}^{TDS}) = \beta_0 + \eta_{ij} + \beta_1 X_{ij}^{age} + \beta_2 X_{ij}^{area} + \beta_3 X_i^{ethn} + \beta_4 X_i^{gender} + \beta_5 X_{ij}^{nattyp} + \beta_6 X_i^{par} + \beta_7 X_i^{schtyp} + \varepsilon_i \tag{5}$$

where $\beta_0$ was the global intercept, $\beta_1, \dots, \beta_7$ were the regression coefficients associated with the covariates, $\varepsilon_i$ was the random effect for child $i$ and $\eta_{ij}$ was the random effect for time $j$ nested in child $i$.

#### Supplementary Methods 2

##### Executive function (EF)

We specified $Y_{ij}^{EF}$ as the measured composite score of three cognitive tests measured during baseline assessment at the schools. As $Y_{ij}^{EF}$ was a continuous variable, and after standardization it can assume any value in $\mathbb{R}$, it was reasonable to assume the following Gaussian distribution with $j = 1, \dots, J = 39$ (total number of schools) and $i = 1, \dots, I =$ 6,386 (total number of children):

$$Y_{ij}^{EF} \sim N(\mu_{ij}^{EF},\ \sigma_{EF}^2) \quad (7)$$

where $\sigma_{EF}^2$ was the variance. We therefore specified a linear model for $\mu_{ij}^{EF}$:

$$\mu_{ij}^{EF} = \beta_0 + \eta_{ij} + \beta_1 X_i^{age} + \beta_2 X_i^{air} + \beta_3 X_i^{area} + \beta_4 X_i^{ethn} + \beta_5 X_i^{gender} + \beta_6 X_i^{nattyp} + \beta_7 X_i^{par} + \beta_8 X_i^{schtyp} \quad (8)$$

where $\beta_0$ was the $EF$ global intercept, $\beta_1, \dots, \beta_8$ were the regression coefficients associated with the covariates and $\eta_{ij}$ was the random effect for school $j$ with child $i$. Air = air pollution; area = area-level deprivation; ethn = ethnicity; nattyp = natural habitat type; par = parental occupation; schtyp = school type.

##### Strengths and Difficulties Questionnaire (SDQ) total difficulties score

We modelled our outcome SDQ total difficulties score with a Poisson distribution. We started by specifying $Y_{ij}^{TDS}$ as the observed number of behavioral difficulties with $j = 1, \dots, J = 39$ (total number of schools) and $i = 1, \dots, I =$ 6,386 (total number of children), and treated these variables as count data to specify the Poisson model:

$$Y_{ij}^{TDS} \sim Poisson\ (\lambda_{ij}^{TDS}\ E_{ij}^{TDS}) \quad (9)$$

where $E_{ij}^{TDS}$ represented the expected number behavioral difficulties and $\lambda_{ij}^{TDS}$ represented the relative risk of behavioral difficulties. We therefore specified a regression model on the log link transformed $\lambda_{ij}^{TDS}$:

$$\log\left(\lambda_{ij}^{TDS}\right) = \beta_0 + \eta_{ij} + \beta_1 X_i^{age} + \beta_2 X_i^{area} + \beta_3 X_i^{ethn} + \beta_4 X_i^{gender} + \beta_5 X_i^{nattyp} + \beta_6 X_i^{par} + \beta_7 X_i^{schtyp} + \varepsilon_i \quad (10)$$

where $\beta_0$ is the global intercept, $\beta_1, \dots, \beta_7$ were the regression coefficients associated with the covariates, $\eta_{ij}$ was the random effect for school $j$ with child $i$, and $\varepsilon_i$ was the random effect for child $i$. We included an additional random effect $\varepsilon_i \sim N(0, \sigma_\varepsilon^2)$ for child $i$ to account for overdispersion, which is typically present when using a Poisson model[2].

**KIDSCREEN-10 Questionnaire Health-Related Quality of Life (HRQoL) score**

We modelled our binary outcome KIDSCREEN-10 Questionnaire HRQoL score with a Binomial distribution with $j = 1, \dots, J = 39$ (total number of schools) and $i = 1, \dots, I =$ 6,386 (total number of children). We model the probability of low overall wellbeing $p_{ij}$ of child $i$ at school $j$ using the logit link function:

$$\text{logit}(p_{ij}) = lo\,g\left(\frac{p_{ij}}{1-p_{ij}}\right) = \beta_0 + \eta_{ij} + \beta_1 X_i^{age} + \beta_2 X_i^{area} + \beta_3 X_i^{ethn} + \beta_4 X_i^{gender} + \beta_5 X_i^{nattyp} + \beta_6 X_i^{par} + \beta_7 X_i^{schtyp} \quad (11)$$

where $\beta_0$ was the global intercept, $\beta_1, \dots, \beta_7$ were the regression coefficients associated with the covariates and $\eta_{ij}$ was the random effect for school $j$ with child $i$.

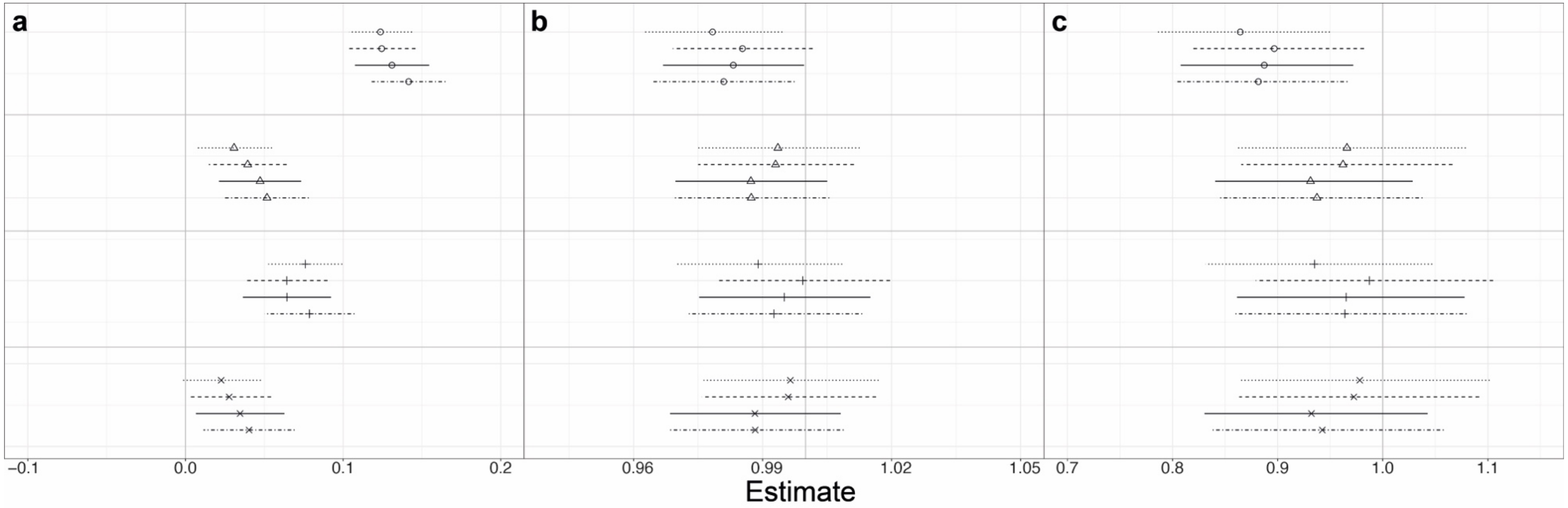


**Supplementary Figure 1. Comparison of different buffer areas to investigate the association between natural space daily exposure rates (DER) and cognitive development and mental health from late childhood to early adolescence.** The association between (a) executive function (EF) score, (b) Strengths and Difficulties Questionnaire (SDQ) total difficulties score (TDS) and (c) KIDSCREEN-10 Questionnaire Health-Related Quality of Life (HRQoL) score and with natural space DER in buffer areas of 50 m (dotted line), 100 m (dashed line), 250 m (solid line) and 500 m (dotdash line) around the home and school area. Four models were fitted: (○) unadjusted (△) adjusted for the effect of ethnicity and school type, (+) adjusted for socioeconomic factors which includes parental occupation and area-level deprivation and (×) adjusted for all risk factors which includes ethnicity, school type, parental occupation and area-level deprivation. All four models were adjusted for age and gender, in the case of EF additionally adjusted for air pollution, and plotted with posterior mean and 95% credible intervals (CI). The vertical line (in grey) is the reference line and significance can be deduced when the 95% CI excludes zero for the EF, and excludes one for the SDQ TDS and HRQoL score.

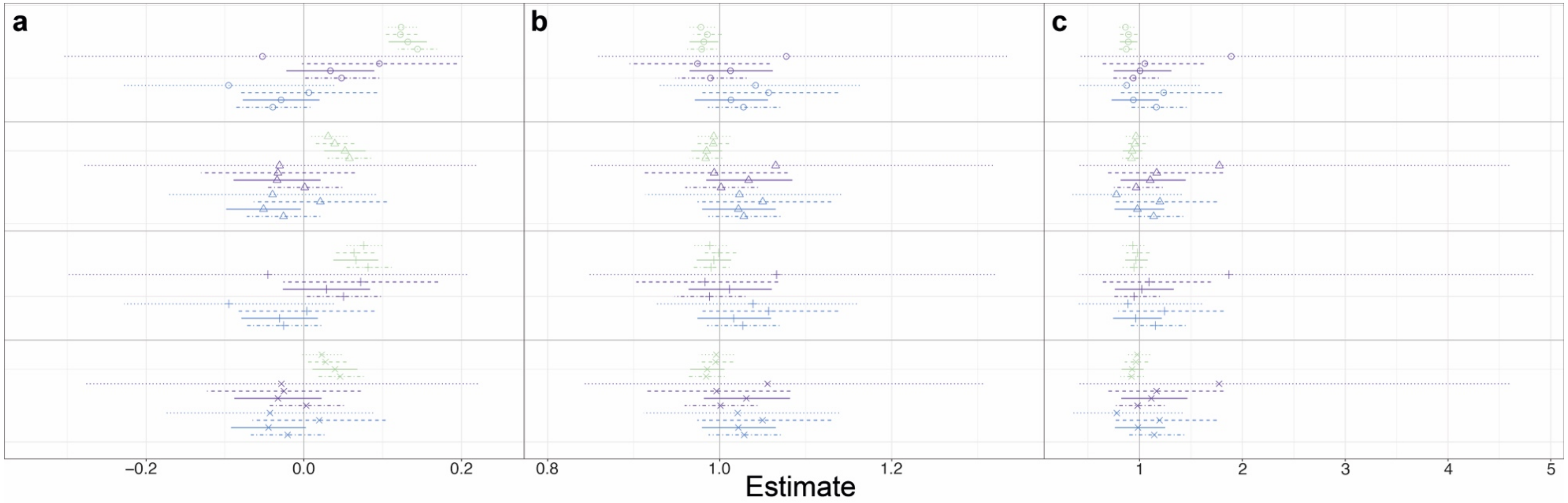


**Supplementary Figure 2. Comparison of different buffer areas to investigate the association between green and blue space daily exposure rates (DER) and cognitive development and mental health from late childhood to early adolescence.** The association between (a) executive function (EF) score, (b) Strengths and Difficulties Questionnaire (SDQ) total difficulties score (TDS) and (c) KIDSCREEN-10 Questionnaire Health-Related Quality of Life (HRQoL) score with the DER of green space (—●—), blue space level 2 (—●—) and blue space level 3 (—●—) in buffer areas of 50 m (dotted line), 100 m (dashed line), 250 m (solid line) and 500 m (dotdash line) around the home and school area. Four models were fitted: (○) unadjusted (△) adjusted for the effect of ethnicity and school type, (+) adjusted for socioeconomic factors which includes parental occupation and area-level deprivation and (×) adjusted for all risk factors which includes ethnicity, school type, parental occupation and area-level deprivation. All four models were adjusted for age and gender, in the case of EF additionally adjusted for air pollution, and plotted with posterior mean and 95% credible intervals (CI). The vertical line (in grey) is the reference line and significance can be deduced when the 95% CI excludes zero for the EF, and excludes one for the SDQ TDS and HRQoL score.

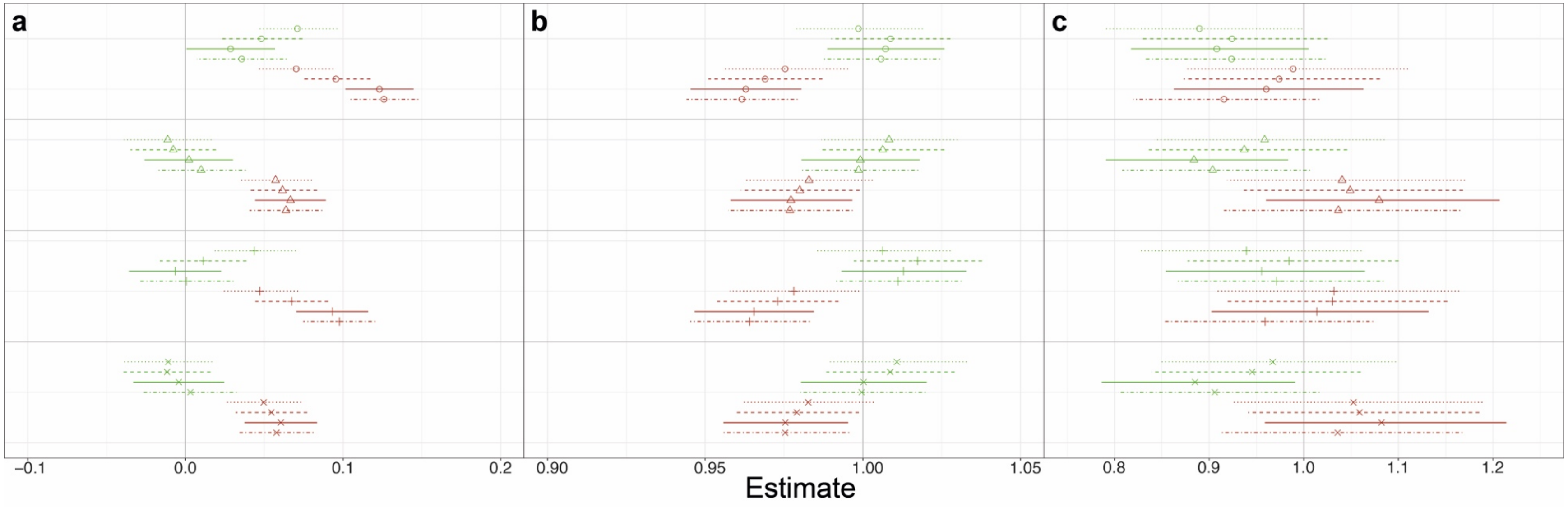


**Supplementary Figure 3. Comparison of different buffer areas to investigate the association between grassland and woodland daily exposure rates (DER) and cognitive development and mental health from late childhood to early adolescence.** The association between (a) executive function (EF) score, (b) Strengths and Difficulties Questionnaire (SDQ) total difficulties score (TDS) and (c) KIDSCREEN-10 Questionnaire Health-Related Quality of Life (HRQoL) score with the DER of grassland (—●—) and woodland (—●—) in buffer areas of 50 m (dotted line), 100 m (dashed line), 250 m (solid line) and 500 m (dotdash line) around the home and school area. Four models were fitted: (○) unadjusted (△) adjusted for the effect of ethnicity and school type, (+) adjusted for socioeconomic factors which includes parental occupation and area-level deprivation and (×) adjusted for all risk factors which includes ethnicity, school type, parental occupation and area-level deprivation. All four models were adjusted for age and gender, in the case of EF additionally adjusted for air pollution, and plotted with posterior mean and 95% credible intervals (CI). The vertical line (in grey) is the reference line and significance can be deduced when the 95% CI excludes zero for the EF, and excludes one for the SDQ TDS and HRQoL score.

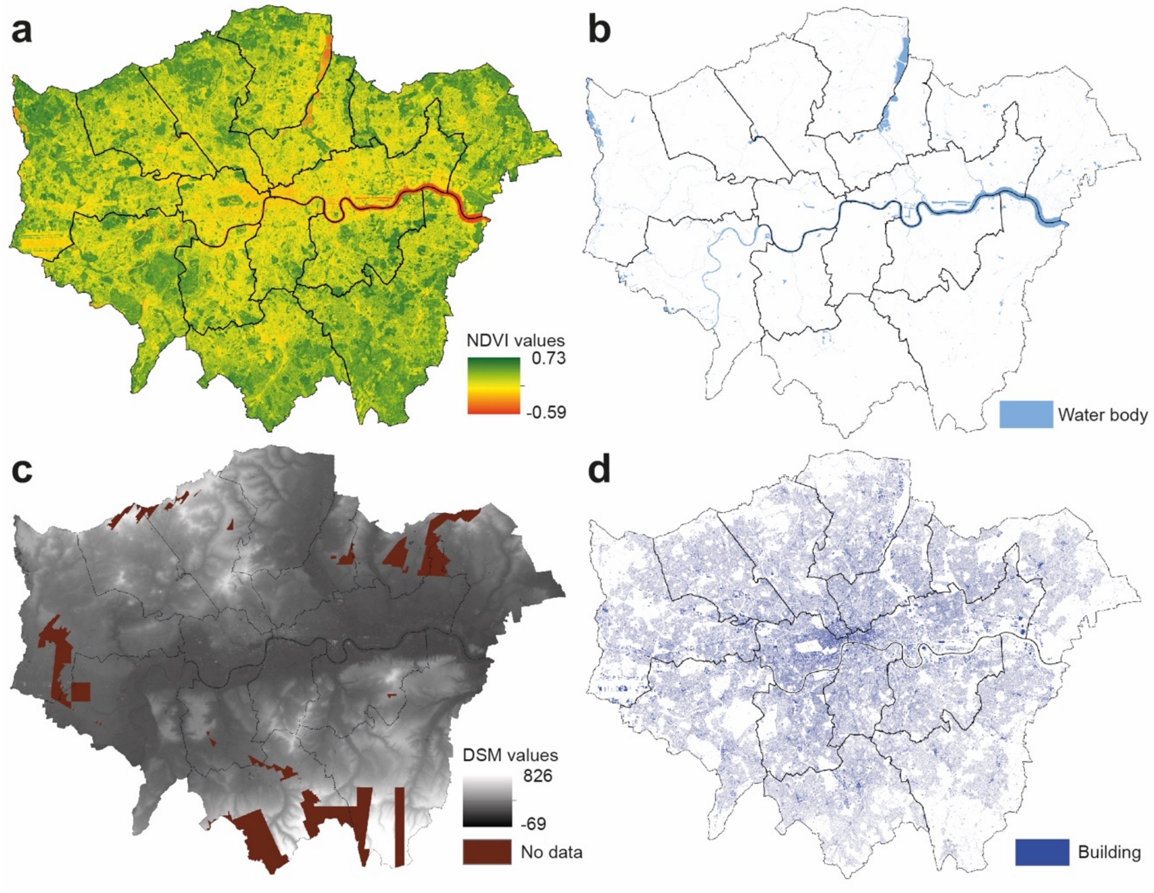


**Supplementary Figure 4. Environmental datasets used to quantify natural habitat exposure.** (a) Normalized Difference Vegetation Index, (b) combined surface and tidal water body map, (c) airborne Light Detection and Ranging map of the Digital Surface Model and (d) buildings map (all images in this figure were restricted to the area of Greater London for visualization purposes).

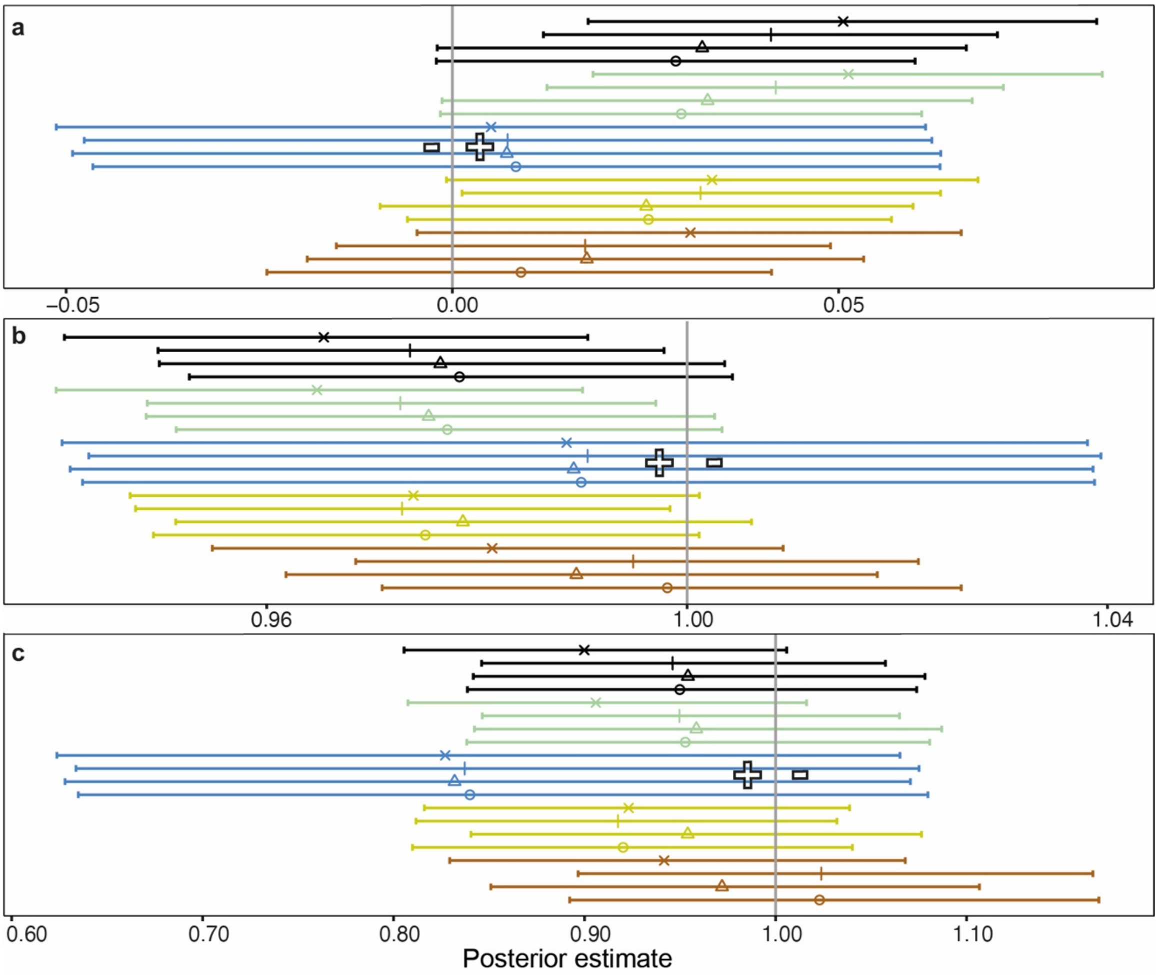


**Supplementary Figure 5. Cross-sectional analysis of the associations between our environmental daily exposure rates (DER), cognitive performance and mental health across London.** The association between the (a) executive function (EF) score, (b) Strengths and Difficulties Questionnaire total difficulties score and (c) KIDSCREEN-10 Questionnaire Health-Related Quality of Life score with the DER of natural space (-●-), green space (-●-), blue space level 3 (-●-), grassland (-●-) and woodland (-●-). We only represented blue space level 3 in this figure. Four models were fitted: (○) unadjusted (△) adjusted for the effect of ethnicity and school type, (+) adjusted for socioeconomic factors which includes parental occupation and area-level deprivation and (×) adjusted for all risk factors which includes ethnicity, school type, parental occupation and area-level deprivation. All four models were adjusted for age and gender, plotted with 95% credible intervals (CI), and models with EF as the outcome were additionally adjusted for air pollution. The vertical line (in grey) is the reference line and is set to zero or one depending on the probability distribution used in each model (Supplementary Methods 2). Hollow plus or minus sign indicated whether the association was positive or negative for cognitive performance or mental health.

**Supplementary Table 1. Median (Q1, Q3) and Pearson's correlation coefficient between estimates of natural habitat daily exposure rate (DER).**

| | *n* | Median (Q1, Q3) | Natural space DER | Green space DER | Blue space DER | | | Grassland DER | Woodland DER |
|---|---|---|---|---|---|---|---|---|---|
| | | | | | Level 1 (ref) | Level 2 | Level 3 | | |
| Natural space DER | 3,563 | 0.53 (0.37, 0.67) | 1 | 0.99 | - | - | - | 0.94 | 0.63 |
| Green space DER | 3,563 | 0.53 (0.36, 0.67) | | 1 | - | - | - | 0.95 | 0.64 |
| Blue space DER | | | | | | | | | |
| Level 1 (ref) | 2,383 | - | | | 1 | - | - | - | - |
| Level 2 | 473 | - | | | | 1 | - | - | - |
| Level 3 | 707 | - | | | | | 1 | - | - |
| Grassland DER | 3,367 | 0.38 (0.25, 0.49) | | | | | | 1 | 0.38 |
| Woodland DER | 3,367 | 0.06 (0.04, 0.11) | | | | | | | 1 |

**Supplementary Table 2. Comparison of fully adjusted models with the executive function (EF) and environmental daily exposure rates (DER) based on daytime (12 hrs) or full day (24 hrs) weighting.** Association between our environmental DER and EF in a buffer area of 250 m. We applied a different weighting on the proportionate presence of each environmental DER based on daytime (12 hrs) and a full day (24 hrs). We fully adjusted all models for age, air pollution, area-level deprivation, ethnicity, gender, parental occupation and school type. Model I (M I) contained natural space DER, Model II (M II) contained green and blue space DER and Model III (M III) contained grassland and woodland DER. Significance was indicated with an asterisk (*) and can be deduced when the 95% credible interval (CI) excluded zero for these models. Qn1, Qn2, Qn3, Qn4 and Qn5 represented the first, second, third, fourth and fifth quintiles of the Carstairs deprivation index, respectively; occ=occupations; emp=employers.

| | Daytime weighting (12 hrs) | | | Full day weighting (24 hrs) | | |
|---|---|---|---|---|---|---|
| | M I: Posterior mean (95% CI) | M II: Posterior mean (95% CI) | M III: Posterior mean (95% CI) | M I: Posterior mean (95% CI) | MII: Posterior mean (95% CI) | M III: Posterior mean (95% CI) |
| α (intercept) | 0.33 (0.27, 0.39)* | 0.35 (0.29, 0.41)* | 0.31 (0.25, 0.37)* | 0.27 (0.21, 0.33)* | 0.29 (0.23, 0.35)* | 0.25 (0.19, 0.31)* |
| Natural space DER | 0.03 (0.006, 0.06)* | - | - | 0.02 (-0.001, 0.04)* | - | - |
| Green space DER | - | 0.03 (0.01, 0.06)* | - | - | 0.02 (0.01, 0.04)* | - |
| Blue space DER | | | | | | |
| Level 1 (ref) | - | 0 (ref) | - | - | 0 (ref) | - |
| Level 2 | - | -0.03 (-0.08, 0.02) | - | - | -0.04 (-0.10, 0.01) | - |
| Level 3 | - | -0.04 (-0.09, 0.01) | - | - | -0.02 (-0.07, 0.01) | - |
| Grassland DER | - | - | -0.01 (-0.03, 0.02) | - | - | 0.01 (-0.02, 0.02) |
| Woodland DER | - | - | 0.06 (0.03, 0.08)* | - | - | 0.04 (0.02, 0.06)* |
| Parental occupation | | | | | | |
| Managerial/professional occ. | 0 (ref) | 0 (ref) | 0 (ref) | 0 (ref) | 0 (ref) | 0 (ref) |
| Intermediate occ. | 0.01 (-0.04, 0.06) | 0.01 (-0.04, 0.06) | 0.01 (-0.04, 0.06) | 0.03 (-0.02, 0.08) | 0.02 (-0.02, 0.08) | 0.03 (-0.02, 0.08) |
| Small emp./own-account workers | -0.01 (-0.05, 0.03) | -0.01 (-0.05, 0.03) | -0.01 (-0.05, 0.04) | -0.01 (-0.06, 0.03) | -0.01 (-0.05, 0.03) | -0.01 (-0.05, 0.03) |
| Lower supervisory/technical occ. | -0.09 (-0.16, -0.01)* | -0.09 (-0.10, -0.01)* | -0.08 (-0.16, -0.01)* | -0.07 (-0.15, -0.01)* | -0.07 (-0.15, -0.01)* | -0.07 (-0.15, -0.01)* |
| Semi-routine/routine occ. | -0.03 (-0.08, 0.02) | -0.03 (-0.08, 0.02) | -0.02 (-0.08, 0.02) | -0.01 (-0.05, 0.04) | -0.01 (-0.05, 0.04) | -0.01 (-0.05, 0.04) |
| Area-level deprivation | | | | | | |
| Least deprived (Qn1) | 0 (ref) | 0 (ref) | 0 (ref) | 0 (ref) | 0 (ref) | 0 (ref) |
| Qn2 | 0.05 (-0.01, 0.11) | 0.05 (-0.01, 0.11) | 0.04 (-0.01, 0.11) | 0.04 (-0.01, 0.10) | 0.04 (-0.01, 0.10) | 0.04 (-0.02, 0.10) |
| Qn3 | 0.03 (-0.03, 0.09) | 0.03 (-0.03, 0.09) | 0.02 (-0.04, 0.08) | 0.01 (-0.05, 0.08) | 0.01 (-0.04, 0.08) | 0.01 (-0.05, 0.08) |
| Qn4 | -0.01 (-0.07, 0.05) | -0.01 (-0.07, 0.05) | -0.01 (-0.07, 0.05) | -0.02 (-0.09, 0.04) | -0.02 (-0.09, 0.04) | -0.02 (-0.09, 0.04) |
| Most deprived (Qn5) | -0.03 (-0.10, 0.04) | -0.02 (-0.10, 0.04) | -0.02 (-0.09, 0.04) | -0.04 (-0.12, 0.02) | -0.04 (-0.11, 0.02) | -0.03 (-0.10, 0.03) |
| Gender | | | | | | |
| Male | 0 (ref) | 0 (ref) | 0 (ref) | 0 (ref) | 0 (ref) | 0 (ref) |
| Female | 0.15 (0.11, 0.19)* | 0.15 (0.11, 0.19)* | 0.16 (0.12, 0.19)* | 0.14 (0.11, 0.18)* | 0.14 (0.10, 0.18)* | 0.15 (0.11, 0.19)* |
| Age | 0.02 (0.01, 0.04)* | 0.02 (0.01, 0.04)* | 0.02 (0.01, 0.04)* | 0.02 (0.01, 0.04)* | 0.02 (0.01, 0.04)* | 0.02 (0.01, 0.04)* |
| $NO_2$ DER | 0.03 (0.01, 0.06)* | 0.03 (0.01, 0.06)* | 0.01 (-0.01, 0.04) | 0.005 (0.004, 0.01)* | 0.005 (0.004, 0.01)* | 0.005 (0.004, 0.01)* |
| Ethnicity | | | | | | |
| White | 0 (ref) | 0 (ref) | 0 (ref) | 0 (ref) | 0 (ref) | 0 (ref) |
| Black | -0.15 (-0.21, -0.09)* | -0.15 (-0.21, -0.09)* | -0.15 (-0.21, -0.09)* | -0.15 (-0.21, -0.09)* | -0.15 (-0.21, -0.09)* | -0.15 (-0.21, -0.09)* |
| Asian | 0.07 (0.02, 0.12)* | 0.06 (0.02, 0.11)* | 0.06 (0.01, 0.11)* | 0.06 (0.02, 0.11)* | 0.06 (0.01, 0.11)* | 0.06 (0.01, 0.11)* |
| Mixed | 0.01 (-0.04, 0.08) | 0.01 (-0.04, 0.08) | 0.01 (-0.05, 0.07) | 0.01 (-0.04, 0.08) | 0.01 (-0.04, 0.08) | 0.01 (-0.05, 0.07) |

| | | | | | | |
|---|---|---|---|---|---|---|
| Other | -0.12 (-0.32, 0.08) | -0.12 (-0.32, 0.08) | -0.11 (-0.31, 0.08) | -0.11 (-0.32, 0.08) | -0.11 (-0.32, 0.08) | -0.12 (-0.32, 0.08) |
| School type | | | | | | |
| Independent | 0 (ref) | 0 (ref) | 0 (ref) | 0 (ref) | 0 (ref) | 0 (ref) |
| State | -0.32 (-0.38, -0.27)* | -0.33 (-0.38, -0.28)* | -0.30 (-0.35, -0.25)* | -0.35 (-0.40, -0.30)* | -0.36 (-0.41, -0.31)* | -0.32 (-0.38, -0.27)* |

**Supplementary Table 3. Comparison of fully adjusted models with Strengths and Difficulties Questionnaire (SDQ) total difficulties score (TDS) and environmental daily exposure rates (DER) based on daytime (12 hrs) or full day (24 hrs) weighting.** Association between our environmental DER and SDQ TDS in a buffer area of 250 m. We applied a different weighting on the proportionate presence of each environmental DER based on daytime (12 hrs) and a full day (24 hrs). We fully adjusted all models for age, area-level deprivation, ethnicity, gender, parental occupation and school type. Model I (M I) contained natural space DER, Model II (M II) contained green and blue space DER and Model III (M III) contained grassland and woodland DER. Significance was indicated with an asterisk (*) and can be deduced when the 95% credible interval (CI) excluded one for these models. Qn1, Qn2, Qn3, Qn4 and Qn5 represented the first, second, third, fourth and fifth quintiles of the Carstairs deprivation index, respectively; occ=occupations; emp=employers.

| | Daytime weighting (12 hrs) | | | Full day weighting (24 hrs) | | |
|---|---|---|---|---|---|---|
| | M I: Posterior mean (95% CI) | M II: Posterior mean (95% CI) | M III: Posterior mean (95% CI) | M I: Posterior mean (95% CI) | M II: Posterior mean (95% CI) | M III: Posterior mean (95% CI) |
| α (intercept) | 8.44 (8.01, 8.87)* | 8.33 (7.89, 8.79)* | 8.51 (8.09, 8.95)* | 8.48 (8.04, 8.94)* | 8.38 (7.92, 8.85)* | 8.54 (8.10, 9)* |
| Natural space DER | 0.98 (0.96, 1.01) | - | - | 0.98 (0.96, 1.01) | - | - |
| Green space DER | - | 0.98 (0.96, 1.01) | - | - | 0.98 (0.96, 1.01) | - |
| Blue space DER | | | | | | |
| Level 1 (ref) | - | 1 (ref) | - | - | 1 (ref) | - |
| Level 2 | - | 1.03 (0.98, 1.08) | - | - | 1.01 (0.96, 1.06) | - |
| Level 3 | - | 1.02 (0.97, 1.06) | - | - | 1.03 (0.99, 1.07) | - |
| Grassland DER | - | - | 1 (0.98, 1.02) | - | - | 0.99 (0.97, 1.01) |
| Woodland DER | - | - | 0.97 (0.95, 0.99)* | - | - | 0.97 (0.95, 0.99)* |
| Parental occupation | | | | | | |
| Managerial/professional occ. | 1 (ref) | 1 (ref) | 1 (ref) | 1 (ref) | 1 (ref) | 1 (ref) |
| Intermediate occ. | 0.93 (0.89, 0.98)* | 0.93 (0.89, 0.98)* | 0.93 (0.88, 0.98)* | 0.93 (0.89, 0.98) | 0.93 (0.89, 0.98)* | 0.93 (0.88, 0.98)* |
| Small emp./own-account workers | 0.99 (0.95, 1.03) | 0.99 (0.95, 1.03) | 0.99 (0.95, 1.03) | 0.99 (0.95, 1.03) | 0.99 (0.95, 1.03) | 0.99 (0.95, 1.03) |
| Lower supervisory/technical occ. | 1 (0.93, 1.07) | 1 (0.93, 1.07) | 0.99 (0.92, 1.06) | 1.01 (0.93, 1.07) | 1.01 (0.93, 1.07) | 0.99 (0.93, 1.07) |
| Semi-routine/routine occ. | 0.98 (0.93, 1.02) | 0.98 (0.93, 1.02) | 0.97 (0.93, 1.02) | 0.98 (0.93, 1.03) | 0.98 (0.93, 1.02) | 0.98 (0.93, 1.02) |
| Area-level deprivation | | | | | | |
| Least deprived (Qn1) | 1 (ref) | 1 (ref) | 1 (ref) | 1 (ref) | 1 (ref) | 1 (ref) |
| Qn2 | 0.99 (0.94, 1.05) | 0.99 (0.94, 1.05) | 1.01 (0.94, 1.06) | 0.99 (0.93, 1.05) | 0.99 (0.94, 1.05) | 0.99 (0.94, 1.05) |
| Qn3 | 1.03 (0.97, 1.09) | 1.02 (0.96, 1.09) | 1.03 (0.97, 1.09) | 1.02 (0.96, 1.08) | 1.02 (0.96, 1.08) | 1.02 (0.96, 1.09) |
| Qn4 | 1.02 (0.96, 1.08) | 1.02 (0.95, 1.08) | 1.02 (0.96, 1.08) | 1.01 (0.95, 1.08) | 1.01 (0.95, 1.08) | 1.01 (0.95, 1.08) |
| Most deprived (Qn5) | 1.01 (0.94, 1.07) | 1.01 (0.94, 1.07) | 1.01 (0.94, 1.07) | 1.01 (0.93, 1.07) | 1.01 (0.93, 1.07) | 1.01 (0.93, 1.07) |
| Gender | | | | | | |
| Male | 1 (ref) | 1 (ref) | 1 (ref) | 1 (ref) | 1 (ref) | 1 (ref) |
| Female | 1.05 (1.02, 1.09)* | 1.05 (1.02, 1.09)* | 1.05 (1.02, 1.09)* | 1.05 (1.02, 1.09)* | 1.05 (1.02, 1.09)* | 1.05 (1.02, 1.09)* |
| Age | 1.01 (0.99, 1.02) | 1.01 (0.99, 1.02) | 1.01 (0.99, 1.02) | 1.01 (0.99, 1.02) | 1.01 (0.99, 1.02)* | 1.01 (0.99, 1.02) |
| Ethnicity | | | | | | |
| White | 1 (ref) | 1 (ref) | 1 (ref) | 1 (ref) | 1 (ref) | 1 (ref) |
| Black | 1.02 (0.97, 1.08) | 1.02 (0.97, 1.08) | 1.02 (0.97, 1.08) | 1.02 (0.97, 1.08) | 1.02 (0.97, 1.08) | 1.02 (0.97, 1.08) |
| Asian | 0.91 (0.87, 0.95)* | 0.91 (0.87, 0.95)* | 0.91 (0.88, 0.96)* | 0.91 (0.87, 0.95)* | 0.91 (0.87, 0.95)* | 0.91 (0.87, 0.95)* |
| Mixed | 1.02 (0.97, 1.08) | 1.02 (0.97, 1.08) | 1.03 (0.97, 1.09) | 1.02 (0.97, 1.08) | 1.02 (0.97, 1.08) | 1.03 (0.97, 1.09) |
| Other | 1.14 (0.96, 1.35) | 1.14 (0.96, 1.35) | 1.14 (0.96, 1.35) | 1.14 (0.96, 1.35) | 1.14 (0.96, 1.35) | 1.14 (0.96, 1.35) |

| | | | | | | |
|---|---|---|---|---|---|---|
| School type | | | | | | |
| Independent | 1 (ref) | 1 (ref) | 1 (ref) | 1 (ref) | 1 (ref) | 1 (ref) |
| State | 1.10 (1.05, 1.15)* | 1.11 (1.06, 1.16)* | 1.09 (1.04, 1.14)* | 1.10 (1.05, 1.15)* | 1.10 (1.05, 1.15)* | 1.09 (1.04, 1.14)* |

**Supplementary Table 4. Comparison of fully adjusted models with KIDSCREEN-10 Questionnaire Health-Related Quality of Life (HRQoL) score and environmental daily exposure rates (DER) based on daytime (12 hrs) or full day (24 hrs) weighting.** Association between our environmental DER and HRQoL score in a buffer area of 250 m. We applied a different weighting on the proportionate presence of each environmental DER based on daytime (12 hrs) and a full day (24 hrs). We fully adjusted all models for age, area-level deprivation, ethnicity, gender, parental occupation and school type. Model I (M I) contained natural space DER, Model II (M II) contained green and blue space DER and Model III (M III) contained grassland and woodland DER. Significance was indicated with an asterisk (*) and can be deduced when the 95% credible interval (CI) excluded one for these models. Qn1, Qn2, Qn3, Qn4 and Qn5 represented the first, second, third, fourth and fifth quintiles of the Carstairs deprivation index, respectively; occ=occupations; emp=employers.

| | Daytime weighting (12 hrs) | | | Full day weighting (24 hrs) | | |
|---|---|---|---|---|---|---|
| | M I: Posterior mean (95% CI) | M II: Posterior mean (95% CI) | M III: Posterior mean (95% CI) | M I: Posterior mean (95% CI) | M II: Posterior mean (95% CI) | M III: Posterior mean (95% CI) |
| α (intercept) | 0.02 (0.01, 0.04)* | 0.02 (0.01, 0.04)* | 0.02 (0.01, 0.03)* | 0.03 (0.01, 0.04)* | 0.02 (0.01, 0.04)* | 0.02 (0.01, 0.03)* |
| Natural space DER | 0.93 (0.83, 1.04) | - | - | 0.93 (0.83, 1.05) | - | - |
| Green space DER | - | 0.92 (0.82, 1.03) | - | - | 0.93 (0.82, 1.04) | - |
| Blue space DER | | | | | | |
| Level 1 (ref) | - | 1 (ref) | - | - | 1 (ref) | - |
| Level 2 | - | 1.11 (0.82, 1.46) | - | - | 1.11 (0.83, 1.44) | - |
| Level 3 | - | 0.98 (0.76, 1.24) | - | - | 0.98 (0.74, 1.25) | - |
| Grassland DER | - | - | 0.88 (0.78, 0.99)* | - | - | 0.88 (0.78, 0.99)* |
| Woodland DER | - | - | 1.08 (0.95, 1.21) | - | - | 1.10 (0.97, 1.24) |
| Parental occupation | | | | | | |
| Managerial/professional occ. | 1 (ref) | 1 (ref) | 1 (ref) | 1 (ref) | 1 (ref) | 1 (ref) |
| Intermediate occ. | 0.83 (0.56, 1.17) | 0.83 (0.56, 1.17) | 0.84 (0.56, 1.18) | 0.83 (0.56, 1.17) | 0.83 (0.56, 1.17) | 0.84 (0.56, 1.18) |
| Small emp./own-account workers | 0.99 (0.75, 1.29) | 0.99 (0.75, 1.29) | 1.01 (0.75, 1.30) | 1.01 (0.75, 1.29) | 1.01 (0.75, 1.29) | 1.01 (0.76, 1.31) |
| Lower supervisory/technical occ. | 1.42 (0.90, 2.10) | 1.43 (0.90, 2.11) | 1.44 (0.91, 2.13) | 1.43 (0.90, 2.11) | 1.43 (0.90, 2.11) | 1.44 (0.91, 2.13) |
| Semi-routine/routine occ. | 0.95 (0.67, 1.30) | 0.95 (0.67, 1.30) | 0.96 (0.68, 1.31) | 0.96 (0.68, 1.30) | 0.96 (0.67, 1.30) | 0.97 (0.68, 1.32) |
| Area-level deprivation | | | | | | |
| Least deprived (Qn1) | 1 (ref) | 1 (ref) | 1 (ref) | 1 (ref) | 1 (ref) | 1 (ref) |
| Qn2 | 1.17 (0.80, 1.65) | 1.16 (0.80, 1.64) | 1.16 (0.80, 1.63) | 1.16 (0.80, 1.64) | 1.15 (0.79, 1.63) | 1.16 (0.80, 1.63) |
| Qn3 | 0.99 (0.67, 1.41) | 0.98 (0.66, 1.40) | 0.98 (0.66, 1.39) | 0.98 (0.66, 1.41) | 0.97 (0.65, 1.39) | 0.99 (0.67, 1.41) |
| Qn4 | 0.91 (0.61, 1.31) | 0.90 (0.60, 1.30) | 0.91 (0.61, 1.31) | 0.91 (0.60, 1.31) | 0.90 (0.59, 1.30) | 0.93 (0.62, 1.33) |
| Most deprived (Qn5) | 1.06 (0.69, 1.55) | 1.04 (0.69, 1.53) | 1.06 (0.70, 1.55) | 1.06 (0.69, 1.56) | 1.05 (0.68, 1.54) | 1.10 (0.72, 1.61) |
| Gender | | | | | | |
| Male | 1 (ref) | 1 (ref) | 1 (ref) | 1 (ref) | 1 (ref) | 1 (ref) |
| Female | 1.95 (1.59, 2.38)* | 1.96 (1.59, 2.39)* | 1.98 (1.61, 2.42)* | 1.95 (1.59, 2.38)* | 1.95 (1.59, 2.39)* | 1.97 (1.60 ,2.40)* |
| Age | 1.09 (0.99, 1.20) | 1.09 (0.99, 1.20) | 1.08 (0.98, 1.19) | 1.10 (1, 1.20)* | 1.10 (1, 1.21)* | 1.09 (0.99, 1.19) |
| Ethnicity | | | | | | |
| White | 1 (ref) | 1 (ref) | 1 (ref) | 1 (ref) | 1 (ref) | 1 (ref) |
| Black | 1.68 (1.24, 2.21)* | 1.67 (1.24, 2.21)* | 1.68 (1.24, 2.21)* | 1.68 (1.24, 2.21)* | 1.67 (1.24, 2.20)* | 1.68 (1.24, 2.22)* |
| Asian | 0.88 (0.67, 1.13) | 0.88 (0.67, 1.13) | 0.86 (0.66, 1.10) | 0.88 (0.67, 1.13) | 0.88 (0.67, 1.13) | 0.87 (0.66, 1.11) |
| Mixed | 1.81 (1.33, 2.40)* | 1.81 (1.33, 2.40)* | 1.78 (1.31, 2.36)* | 1.81 (1.33, 2.40)* | 1.81 (1.33, 2.39)* | 1.78 (1.31, 2.35)* |
| Other | 2.63 (1.06, 5.23)* | 2.65 (1.07, 5.27)* | 2.63 (1.07, 5.23)* | 2.62 (1.06, 5.20)* | 2.64 (1.06, 5.26)* | 2.63 (1.06, 5.23)* |

| | | | | | | |
|---|---|---|---|---|---|---|
| School type | | | | | | |
| Independent | 1 (ref) | 1 (ref) | 1 (ref) | 1 (ref) | 1 (ref) | 1 (ref) |
| State | 1.57 (1.19, 2.04)* | 1.60 (1.21, 2.09)* | 1.71 (1.28, 2.24)* | 1.56 (1.18, 2.03)* | 1.60 (1.20, 2.09)* | 1.69 (1.27, 2.22)* |

**Supplementary Table 5. Contribution of risk factor groups based on the difference in pseudo R-squared between the full fixed-effects only Model I (M I) and M I excluding environmental, demographic or socioeconomic variables.** The full fixed-effect only M I included environmental (i.e. natural space daily exposure rate [DER] and air pollution), demographic (i.e. gender, age and ethnicity) and socioeconomic variables (parental occupation, area-level deprivation and school type). Mean pseudo R-squared was calculated by dividing the mean squared error between predicted and observed values by the variance of the observed values for each fold in a 10-fold cross validation. Standard error (SE) of the mean pseudo R-squared was calculated by dividing the standard deviation by the square root of the number of measurements. We did not calculate a pseudo R-squared for the Health-Related Quality of Life score because the observed value is binomial, making it impossible to measure a pseudo R-squared.

| | Pseudo R-squared | Difference |
|---|---|---|
| EF | Mean (SE) | |
| Full fixed-effects only model | 0.102 (0.006) | - |
| - Environmental variables | 0.101 (0.006) | 0.001 |
| - Demographic variables | 0.08 (0.004) | 0.022 |
| - Socioeconomic variables | 0.062 (0.004) | 0.04 |
| SDQ TDS | | |
| Full fixed-effects only model | 0.019 (0.003) | - |
| - Environmental variables | 0.018 (0.003) | 0.001 |
| - Demographic variables | 0.01 (0.002) | 0.009 |
| - Socioeconomic variables | 0.01 (0.001) | 0.008 |

**Supplementary Table 6. Characteristics of the baseline and follow-up cohort.** Baseline and follow-up data were based on participants who took part in the computer-based assessment. This study used a subset of children (*n* = 3,568) who had a known home address during the baseline and follow-up assessment. Parental occupation is based on the highest National Statistics Socioeconomic Classification (NS-SEC) level (five-group version) of either parent. Qn1, Qn2, Qn3, Qn4 and Qn5 of area-level deprivation represented the first, second, third, fourth and fifth quintile of the Carstairs deprivation index, respectively.

| | Baseline cohort | | Follow-up cohort | | Subset with home address in baseline and follow-up | |
|---|---|---|---|---|---|---|
| | *n* = 6,612 | | *n* = 5,208 | | *n* = 3,568 | |
| | Median | IQR | Median | IQR | Median | IQR |
| Age (years) | 12.06 | 11.78-12.33 | 14.21 | 13.92-14.56 | 12.96 | 12.02-14.22 |
| Parental occupation | *n* | % | *n* | % | *n* | % |
| Managerial/professional occupations | 3270 | 49.45 | 2788 | 53.53 | 2077 | 58.21 |
| Intermediate occupations | 484 | 7.32 | 283 | 5.43 | 292 | 8.18 |
| Small employers/own-account workers | 908 | 13.73 | 752 | 14.43 | 507 | 14.20 |
| Lower supervisory/technical occupations | 272 | 4.11 | 190 | 3.64 | 161 | 4.51 |
| Semi-routine/routine occupations | 693 | 10.48 | 397 | 7.62 | 398 | 11.15 |
| Missing/not interpretable | 985 | 14.89 | 798 | 15.32 | 133 | 3.72 |
| Area-level deprivation | | | | | | |
| Least deprived (Qn1) | 919 | 13.89 | 821 | 15.76 | 580 | 16.25 |
| Qn2 | 944 | 14.27 | 810 | 15.55 | 561 | 15.72 |
| Qn3 | 1122 | 16.96 | 873 | 16.76 | 620 | 17.37 |
| Qn4 | 1389 | 21 | 1050 | 20.16 | 747 | 20.93 |
| Most deprived (Qn5) | 2024 | 30.61 | 1495 | 28.70 | 1058 | 29.65 |
| Missing | 214 | 3.23 | 159 | 3.05 | 2 | 0.05 |
| Gender | | | | | | |
| Female | 3468 | 52.45 | 2823 | 54.20 | 2069 | 57.98 |
| Male | 3144 | 47.54 | 2385 | 45.79 | 1499 | 42.01 |
| Ethnicity | | | | | | |
| White | 2719 | 41.12 | 2265 | 43.49 | 1617 | 45.31 |
| Black | 980 | 14.82 | 739 | 14.18 | 523 | 14.65 |
| Asian | 1715 | 25.93 | 1354 | 25.99 | 959 | 26.87 |
| Mixed | 712 | 10.76 | 498 | 9.56 | 406 | 11.37 |
| Other/not interpretable | 54 | 0.81 | 28 | 0.53 | 31 | 0.86 |
| Missing | 432 | 6.53 | 324 | 6.22 | 32 | 0.89 |
| Type of school | | | | | | |
| State | 5177 | 78.29 | 3918 | 75.23 | 2556 | 71.63 |
| Independent | 1435 | 21.70 | 1290 | 24.76 | 1012 | 28.36 |

**Supplementary Table 7. Median (Q1, Q3) and Pearson's correlation coefficient between estimates of air pollution daily exposure rate (DER).**

| | *n* | Median (Q1, Q3) | $NO_2$ DER | $NO_x$ DER | $PM_{10}$ DER | $PM_{2.5}$ DER |
|---|---|---|---|---|---|---|
| $NO_2$ DER | 3,305 | 35.67 (33.56, 38.26) | 1 | 0.98 | 0.95 | 0.98 |
| $NO_x$ DER | 3,305 | 63.44 (57.57, 70.48) | | 1 | 0.96 | 0.96 |
| $PM_{10}$ DER | 3,305 | 6.93 (5.91, 8.11) | | | 1 | 0.95 |
| $PM_{2.5}$ DER | 3,305 | 13.17 (12.85, 13.50) | | | | 1 |

**Supplementary Table 8. Cross validation results testing different models for the executive function (EF).** We tested Gaussian models with different random effect (RE) structures between EF and natural space daily exposure rate from late childhood to early adolescence. We used model-selection criteria to identify the best model, i.e. the Deviance Information Criterion (DIC), the Log-Pseudo Marginal Likelihood (LPML) and the pseudo R-squared from 10-fold cross validation where a lower DIC and a higher LPML and pseudo-R squared better support the data. We added penalized complexity priors to models with an asterisk (*) because the precision of the model hyperparameters was far too high with the default prior[1]. We used the standard deviation of the residuals of the fixed effects only model to specify a scale for the standard deviation of the random effects.

| | Unadjusted | Adjusted for ethnicity and school type | Adjusted for socioeconomic status | Adjusted for all |
|---|---|---|---|---|
| DIC | | | | |
| No RE | 12247 | 11900 | 12150 | 11894 |
| RE for child id | 9579 | 9479 | 9567 | 9481 |
| RE for school type | 11958 | 11900 | 11952 | 11894 |
| RE for school id | 11785 | 11740 | 11792 | 11748 |
| RE for child id and school id | 9497 | 9469 | 9501 | 9474 |
| RE for time of visit | 12118 | 11771 | 12020 | 11763 |
| *RE for child id and time of visit (2-level nested model) | 6509 | 6451 | 6509 | 6453 |
| *RE for school id, child id and time of visit (3-level nested model) | 6375 | -1927 | 6363 | -36204 |
| LPML | | | | |
| No RE | -6123 | -5950 | -6075 | -5947 |
| RE for child id | -5157 | -5075 | -5142 | -5076 |
| RE for school type | -5979 | -5950 | -5976 | -5947 |
| RE for school id | -5892 | -5870 | -5896 | -5874 |
| RE for child id and school id | -5070 | -5052 | -5074 | -5056 |
| RE for time of visit | -6059 | -5885 | -6010 | -5881 |
| *RE for child id and time of visit (2-level nested model) | -5156 | -5075 | -5141 | -5076 |
| *RE for school id, child id and time of visit (3-level nested model) | -5070 | -5021 | -5073 | 12822 |
| Pseudo R-squared from 10-fold cross validation | | | | |
| No RE | 0.21 | 0.31 | 0.25 | 0.31 |
| RE for child id | 0.89 | 0.88 | 0.89 | 0.88 |
| RE for school type | 0.29 | 0.31 | 0.30 | 0.31 |
| RE for school id | 0.35 | 0.36 | 0.35 | 0.36 |
| RE for child id and school id | 0.88 | 0.88 | 0.88 | 0.88 |
| RE for time of visit | 0.25 | 0.34 | 0.29 | 0.35 |
| *RE for child id and time of visit (2-level nested model) | 0.98 | 0.98 | 0.98 | 0.98 |
| *RE for school id, child id and time of visit (3-level nested model) | 0.98 | 0.99 | 0.98 | 1 |

**Supplementary Table 9. Cross validation results testing different models for the Strengths and Difficulties Questionnaire (SDQ) total difficulties score (TDS).** We tested Poisson models with different random effect (RE) structures between the SDQ TDS and natural space daily exposure rate from late childhood to early adolescence. We used model-selection criteria to identify the best model, i.e. the Deviance Information Criterion (DIC), the Log-Pseudo Marginal Likelihood (LPML) and the pseudo R-squared from 10-fold cross validation where a lower DIC and a higher LPML and pseudo-R squared better support the data.

| | Unadjusted | Adjusted for ethnicity and school type | Adjusted for socioeconomic status | Adjusted for all |
|---|---|---|---|---|
| DIC | | | | |
| No RE | 43009 | 42765 | 42948 | 42748 |
| RE for child id | 35036 | 35033 | 35041 | 35035 |
| RE for school type | 42884 | 42764 | 42865 | 42748 |
| RE for school id | 42527 | 42462 | 42522 | 42456 |
| RE for child id and school id | 35026 | 35025 | 35027 | 35026 |
| RE for time of visit | 42795 | 42542 | 42735 | 42530 |
| RE for child id and time of visit (2-level nested model) | 34555 | 34550 | 34559 | 34553 |
| RE for school id, child id and time of visit (3-level nested model) | 34542 | 34541 | 34545 | 34543 |
| LPML | | | | |
| No RE | -21509 | -21391 | -21486 | -21390 |
| RE for child id | -18439 | -18431 | -18445 | -18435 |
| RE for school type | -21447 | -21391 | -21445 | -21390 |
| RE for school id | -21294 | -21264 | -21299 | -21269 |
| RE for child id and school id | -18424 | -18421 | -18428 | -18424 |
| RE for time of visit | -21402 | -21281 | -21380 | -21282 |
| RE for child id and time of visit (2-level nested model) | -18213 | -18203 | -18218 | -18208 |
| RE for school id, child id and time of visit (3-level nested model) | -18195 | -18192 | -18200 | -18196 |
| Pseudo R-squared from 10-fold cross validation | | | | |
| No RE | 0.05 | 0.13 | 0.08 | 0.13 |
| RE for child id | 0.87 | 0.87 | 0.87 | 0.87 |
| RE for school type | 0.09 | 0.13 | 0.11 | 0.14 |
| RE for school id | 0.18 | 0.19 | 0.18 | 0.20 |
| RE for child id and school id | 0.87 | 0.87 | 0.87 | 0.87 |
| RE for time of visit | 0.12 | 0.17 | 0.13 | 0.17 |
| RE for child id and time of visit (2-level nested model) | 0.96 | 0.96 | 0.96 | 0.96 |
| RE for school id, child id and time of visit (3-level nested model) | 0.96 | 0.96 | 0.96 | 0.96 |

**Supplementary Table 10. Cross validation results testing different models for the KIDSCREEN-10 Questionnaire Health-Related Quality of Life (HRQoL) score.** We tested Binomial models with different random effect (RE) structures between the HRQoL score and natural space daily exposure rate from late childhood to early adolescence. We used model-selection criteria to identify the best model, i.e. the Deviance Information Criterion (DIC) and the Log-Pseudo Marginal Likelihood (LPML) where a lower DIC and a higher LPML better support the data. We did not use 10-fold cross validation because the observed value is binomial, making it impossible to calculate a pseudo R-squared. We added informative gamma priors to models with an asterisk (*) because the precision of model parameters was far too high with the default prior. We set the mean value of the gamma prior to the inverse of the variance of the residuals of the fixed-effects only model.

| | Unadjusted | Adjusted for ethnicity and school type | Adjusted for socioeconomic status | Adjusted for all |
|---|---|---|---|---|
| DIC | | | | |
| No RE | 4013 | 3970 | 4018 | 3980 |
| RE for child id | 3843 | 3805 | 3840 | 3807 |
| *RE for school type | 3996 | 3970 | 4006 | 3979 |
| *RE for school id | 4004 | 3971 | 4013 | 3980 |
| *RE for child id and school id | 3819 | 3789 | 3820 | 3790 |
| *RE for time of visit | 4015 | 3971 | 4020 | 3981 |
| *RE for child id and time of visit (2-level nested model) | 3823 | 3787 | 3820 | 3788 |
| *RE for school id, child id and time of visit (3-level nested model) | 3811 | 3777 | 3819 | 3788 |
| LPML | | | | |
| No RE | -2006 | -1985 | -2009 | -1990 |
| RE for child id | -1934 | -1916 | -1933 | -1919 |
| *RE for school type | -1998 | -1985 | -2003 | -1990 |
| *RE for school id | -2002 | -1985 | -2006 | -1990 |
| *RE for child id and school id | -1922 | -1906 | -1923 | -1909 |
| *RE for time of visit | -2007 | -1985 | -2010 | -1990 |
| *RE for child id and time of visit (2-level nested model) | -1924 | -1906 | -1925 | -1909 |
| *RE for school id, child id and time of visit (3-level nested model) | -1920 | -1904 | -1924 | -1909 |

#### Supplementary Information references


1. Simpson, D., Rue, H., Martins, T. G., Riebler, A. & Sørbye, S. H. Penalising model component complexity: A principled, practical approach to constructing priors. *Stat. Sci.* **32**, 1–28 (2017).
2. Blangiardo, M., Pirani, M., Kanapka, L., Hansell, A. & Fuller, G. A hierarchical modelling approach to assess multi pollutant effects in time-series studies. *PLoS One* **14**, e0212565 (2019).
